# Medium-term effects of SARS-CoV-2 infection on multiple vital organs, exercise capacity, cognition, quality of life and mental health, post-hospital discharge

**DOI:** 10.1101/2020.10.15.20205054

**Authors:** Betty Raman, Mark Philip Cassar, Elizabeth M Tunnicliffe, Nicola Filippini, Ludovica Griffanti, Fidel Alfaro-Almagro, Thomas Okell, Fintan Sheerin, Cheng Xie, Masliza Mahmod, Ferenc E Mózes, Adam J Lewandowski, Eric O Ohuma, David Holdsworth, Hanan Lamlum, Myles J Woodman, Catherine Krasopoulos, Rebecca Mills, Flora A Kennedy McConnell, Chaoyue Wang, Christoph Arthofer, Frederik J Lange, Jesper Andersson, Mark Jenkinson, Charalambos Antoniades, Keith M Channon, Mayooran Shanmuganathan, Vanessa M Ferreira, Stefan K Piechnik, Paul Klenerman, Christopher Brightling, Nick P Talbot, Nayia Petousi, Najib M Rahman, Ling-Pei Ho, Kate Saunders, John R Geddes, Paul J Harrison, Kyle Pattinson, Matthew J Rowland, Brian J Angus, Fergus Gleeson, Michael Pavlides, Ivan Koychev, Karla L Miller, Clare Mackay, Peter Jezzard, Stephen M Smith, Stefan Neubauer

## Abstract

**Background:** The medium-term effects of Coronavirus disease (COVID-19) on multiple organ health, exercise capacity, cognition, quality of life and mental health are poorly understood.

**Methods:** Fifty-eight COVID-19 patients post-hospital discharge and 30 comorbidity-matched controls were prospectively enrolled for multiorgan (brain, lungs, heart, liver and kidneys) magnetic resonance imaging (MRI), spirometry, six-minute walk test, cardiopulmonary exercise test (CPET), quality of life, cognitive and mental health assessments.

**Findings:** At 2-3 months from disease-onset, 64% of patients experienced persistent breathlessness and 55% complained of significant fatigue. On MRI, tissue signal abnormalities were seen in the lungs (60%), heart (26%), liver (10%) and kidneys (29%) of patients. COVID-19 patients also exhibited tissue changes in the thalamus, posterior thalamic radiations and sagittal stratum on brain MRI and demonstrated impaired cognitive performance, specifically in the executive and visuospatial domain relative to controls. Exercise tolerance (maximal oxygen consumption and ventilatory efficiency on CPET) and six-minute walk distance (405±118m vs 517±106m in controls, p<0.0001) were significantly reduced in patients. The extent of extra-pulmonary MRI abnormalities and exercise tolerance correlated with serum markers of ongoing inflammation and severity of acute illness. Patients were more likely to report symptoms of moderate to severe anxiety (35% versus 10%, p=0.012) and depression (39% versus 17%, p=0.036) and a significant impairment in all domains of quality of life compared to controls.

**Interpretation:** A significant proportion of COVID-19 patients discharged from hospital experience ongoing symptoms of breathlessness, fatigue, anxiety, depression and exercise limitation at 2-3 months from disease-onset. Persistent lung and extra-pulmonary organ MRI findings are common. In COVID-19 survivors, chronic inflammation may underlie multiorgan abnormalities and contribute to impaired quality of life.

**Funding:** NIHR Oxford and Oxford Health Biomedical Research Centres, British Heart Foundation Centre for Research Excellence, UKRI, Wellcome Trust, British Heart Foundation.

## Background

The global impact of Coronavirus disease or COVID-19 has been profound with hundreds of thousands of lives lost and millions affected.^1^ Despite the high mortality seen among hospitalised patients, many have survived, with little known about the medium-to-long term effects of COVID-19 after discharge. Although predominantly a respiratory illness, emerging data suggests that multiorgan injury is common, particularly in those with moderate to severe infections.^2-4^

Studies have shown that the brain, heart, gastrointestinal system and the kidneys are particularly vulnerable to injury during the initial phase of illness.^2-4^ The invasive potential and affinity of severe acute respiratory syndrome coronavirus-2 (SARS-CoV-2) for multiple cell lines, suggests that virus-mediated toxicity may play a central role in promoting multi-system damage.^5^ An exaggerated and dysregulated immune response, endothelial damage, thromboinflammation and coagulopathy have also been implicated in causing injury to these organs.^6^

While it is clear that the extent of infection, inflammatory response and physiological reserve (influenced by obesity, age and comorbidities) are important determinants of clinical outcomes during the initial phase, reports of a chronic maladaptive inflammatory syndrome are also emerging. In particular, inflammatory changes in the lungs have been described in convalescing COVID-19 patients, months after discharge from hospital.^7^ Whether extra-pulmonary organs are also susceptible to ongoing inflammation and injury is poorly understood as are its effects on exercise tolerance, cognition, mental health and quality of life.

In a holistic study of survivors of moderate to severe COVID-19 infection, discharged from hospital, at 2-3 months from disease onset, we aimed to investigate the prevalence of persistent multiorgan injury/inflammation and assess the effects of COVID-19 on physical, psychological, and cognitive health and well-being.

## Patients and methods

### Study population

Fifty-eight patients with moderate to severe laboratory-confirmed (SARS-CoV-2 polymerase chain reaction positive) COVID-19, admitted for treatment at the Oxford University Hospitals National Health Service Foundation Trust between 14^th^ March – 25^th^ May 2020, and 30 uninfected controls (SARS-CoV-2 immunoglobulin negative and asymptomatic) group-matched for age, sex, body mass index (BMI) and risk factors (smoking, diabetes and hypertension) from the community (during the same period) were prospectively enrolled in this study **(appendix, Figure 1, p28)**. For further details on inclusion and exclusion criteria, refer to **appendix, p2**. COVID-19 severity on admission was defined as per the World Health Organisation (WHO) interim clinical guidance (**appendix** for definition, **p2**). Patients with severe/end-stage multi-system comorbidities and contraindications to magnetic resonance imaging were excluded.

This study was registered at ClinicalTrials.gov (**NCT04510025)** and approved in the United Kingdom by the North West Preston Research Ethics Committee (**reference 20/NW/0235**).

### Study Procedures

All subjects consented to have comprehensive multiorgan magnetic resonance imaging (MRI) of the brain, lungs, heart, liver, kidneys, six-minute walk (6MWT) test, spirometry, cardiopulmonary exercise test (CPET), series of questionnaires, and blood tests.

### Multiorgan MRI protocol

A 70 minute multiorgan MRI scan was carried out at 3 Tesla (Prisma, Siemens Healthineers, Erlangen, Germany). Brain MRI included T_1_ weighted imaging, T_2_-Fluid attenuated inversion recovery (FLAIR) to assess (e.g.) inflammation, diffusion-weighted imaging (DWI) to assess ischaemic injury, susceptibility-weighted imaging (SWI) to assess (e.g.) haemorrhage, and quantitative multi-inversion-delay arterial spin labelling (ASL) to assess cerebral blood flow. Lung imaging included a T_2_-weighted half-Fourier-acquisition single-shot turbo spin-echo (HASTE) to qualitatively assess the extent of lung parenchymal involvement. Cardiac assessment included cine imaging, T_1_ and T_2_ mapping, post-contrast T_1_ mapping and late gadolinium enhancement (LGE) imaging to assess biventricular volumes, function, myocardial oedema, diffuse and focal/patchy fibrosis. Liver imaging consisted of a single slice T_1_ map and multiecho gradient echo IDEAL, to quantify fibro-inflammation (T_1_), fat (proton density fat fraction, PDFF) and iron (T_2_*). Multiparametric renal imaging was also undertaken and consisted of a single coronal oblique slice T_1_ map to quantify fibroinflammat ion and an R2* map for renal oxygenation assessment (details in **appendix, p2-5**).^8^

### MRI Image analysis

All images were analysed quantitatively and qualitatively (by expert radiologist, neuroradiologist, cardiac MRI specialists, neuroimage analysts and physicists) in a blinded fashion. Lung MRIs were qualitatively assessed for parenchymal involvement by an expert radiologist. Extent of lung parenchymal abnormalities was scored as 0 (0%), 1 (1–25%), 2 (26–50%), 3 (51–75%), or 4 (76– 100%), respectively. Brain image processing was carried out using an adapted version of the processing pipeline created for the United Kingdom (UK) Biobank brain imaging analysis (https://www.fmrib.ox.ac.uk/ukbiobank/), based around tools from the Oxford Centre for Functional Magnetic Resonance Imaging of the Brain (FMRIB) software library [FSL (https://fsl.fmrib.ox.ac.uk/fsl)] (**appendix, p3,4)**. Assessment of cardiac volumes, function, myocardial T_1_ maps, T_2_ maps, post-contrast T_1_ maps and late gadolinium enhanced images was undertaken using cvi42 software (Circle Cardiovascular Imaging Inc., Version 5.10.1, Calgary, Canada) **(appendix, p4)**. Quantitative analyses of T_1_ and T_2_* for liver, spleen and renal images were carried out as described in the appendix **(appendix, p5)**, including iron-correction for liver T1 with an algorithm related to Liver*MultiScan* cT1 (Perspectum, Oxford) but lacking cross-scanner standardisation and therefore not comparable to LiverMultiScan cT1.

### Spirometry

Spirometry, including forced vital capacity (FVC), and forced expiratory volume at the first second of exhalation (FEV1), was performed as per recommended guidance (**appendix, p5**).

### CPET

Symptom-limited CPET was undertaken using a cycle ergometer. Following two minutes of unloaded cycling, the work rate was increased to 20W, followed by a 10W/min ramp. Participants were encouraged to reach their maximal work rate (**appendix, p6**).

### Six-minute walk test

Participants were asked to walk for six-minutes in a pre-marked corridor. Borg scale, heart rate and oxygen saturation were measured immediately before and after the test (**appendix, p5**).

### Questionnaires

Questionnaires were completed using an electronic data capture (CASTOR EDC, https://www.castoredc.com). Depression, anxiety and quality of life measures were assessed using the Patient Health Questionnaire (PHQ) depression module (PHQ-9)^9^, General Anxiety Disorder Questionnaire (GAD-7)^10^ and Short Form-36 (SF-36) survey^11^. Cognitive function was assessed using the Montreal Cognitive Asessment (MoCA). The Medical Research Council (MRC) dyspnea^12^ scale, Dyspnoea-12 score^13^ and Fatigue Severity Scale (FSS)^14^ were used to assess the extent of breathlessness and fatigue, respectively (**appendix, p6**).

### Laboratory assessments

Blood-based testing consisted of a complete blood count, biochemical analysis, coagulation testing, assessment of liver and renal function, markers of cardiac injury and measures of electrolytes, C-reactive protein (CRP), procalcitonin, and lactate dehydrogenase.

### Admission data collection and blood tests

Details on clinical symptoms or signs, vitals and laboratory findings during admission were extracted from electronic medical records. The severity of disease during hospital admission was graded as per the WHO ordinal scale for clinical improvement (**appendix** for definition, **p2**).

### Statistical analysis

Continuous variables were described using mean and standard deviation (SD) for parametric data and median with interquartile range (IQR) for non-parametric data in the tables. Normality was assessed by visual inspection of histograms. Mean differences between two groups were evaluated using Student’s t-tests or Mann-Whitney U-tests as appropriate. Categorical variables were reported as frequency and percentages. Associations between two groups were compared using the Chi-square test or Fisher’s exact test as appropriate. Pearson’s or Spearman’s correlation coefficients were used to describe the relationship between two variables where relevant. Analyses of brain imaging derived phenotypes (IDP’s) were undertaken after Gaussianisation (quantile normalisation) of all continuous variables and adjusting for age, sex, BMI, diastolic and systolic blood pressure, smoking and head size. The conventional level of statistical significance of 5% was used and not corrected for multiple comparisons. Statistical analyses were performed using SPSS Version 26.0 (IBM, Armonk, NY, USA).

## Results

Mean age of the patient group was 55±13 years and 34/58 (59%) were men (**Table 1**). Thirteen (22%) belonged to Black, Asian and Minority Ethnic groups. Twenty one patients (36%) had required high dependency unit (HDU) or intensive care unit (ICU) admission; 20/21 required mechanical ventilation (non-invasive ventilation or intubation). Median duration of hospitalisation was 8·5 days (IQR 5·0 - 17·0). Patients were assessed between 2-3 months from disease-onset at median interval of 2·3 months (IQR 2·06 - 2·53). Baseline characteristics of patients and group-matched controls are listed in **Table 1**. At follow-up, COVID-19 patients had a mildly increased resting heart rate, respiratory rate and reduced oxygen saturation relative to controls.

**Table 1.**
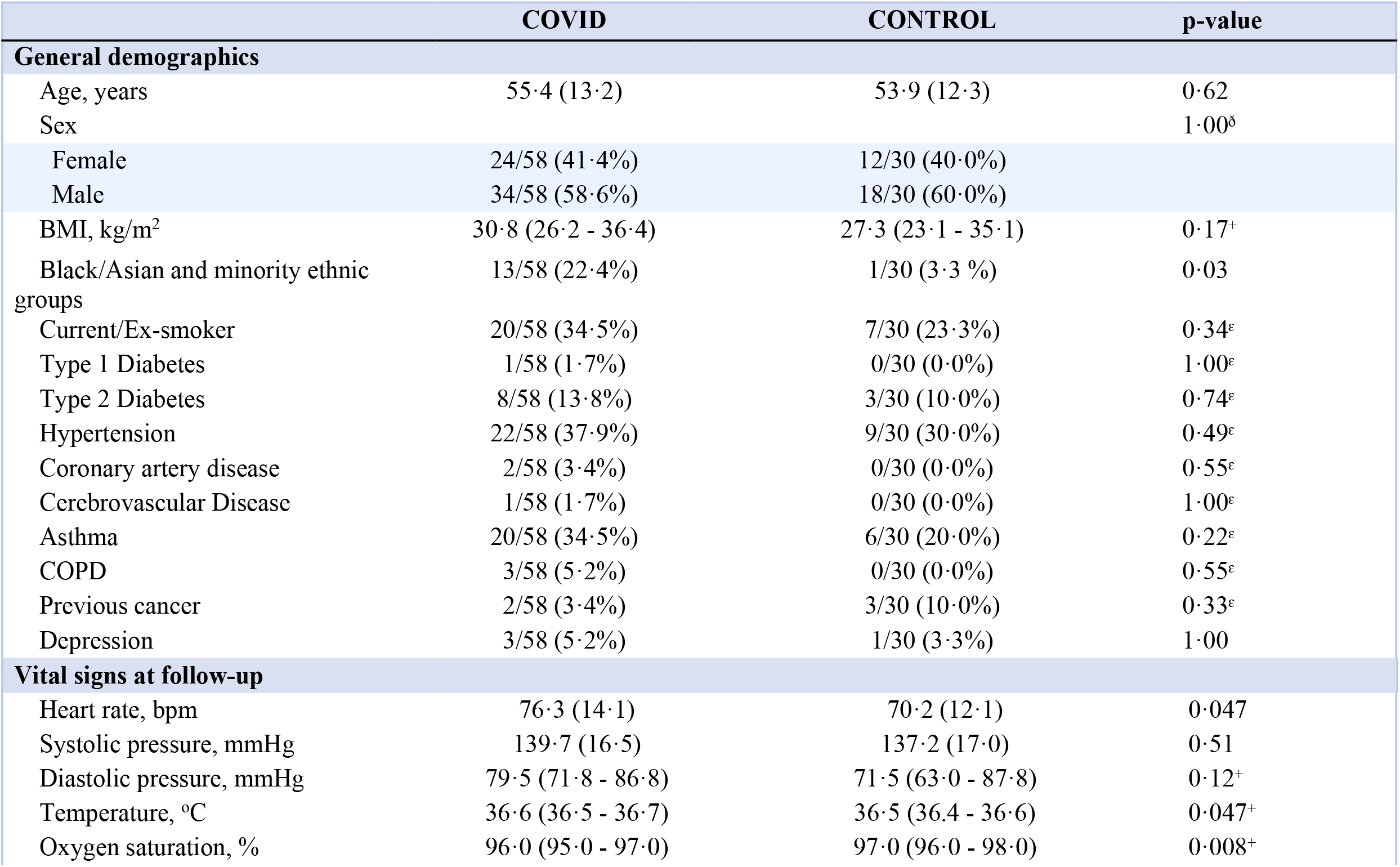
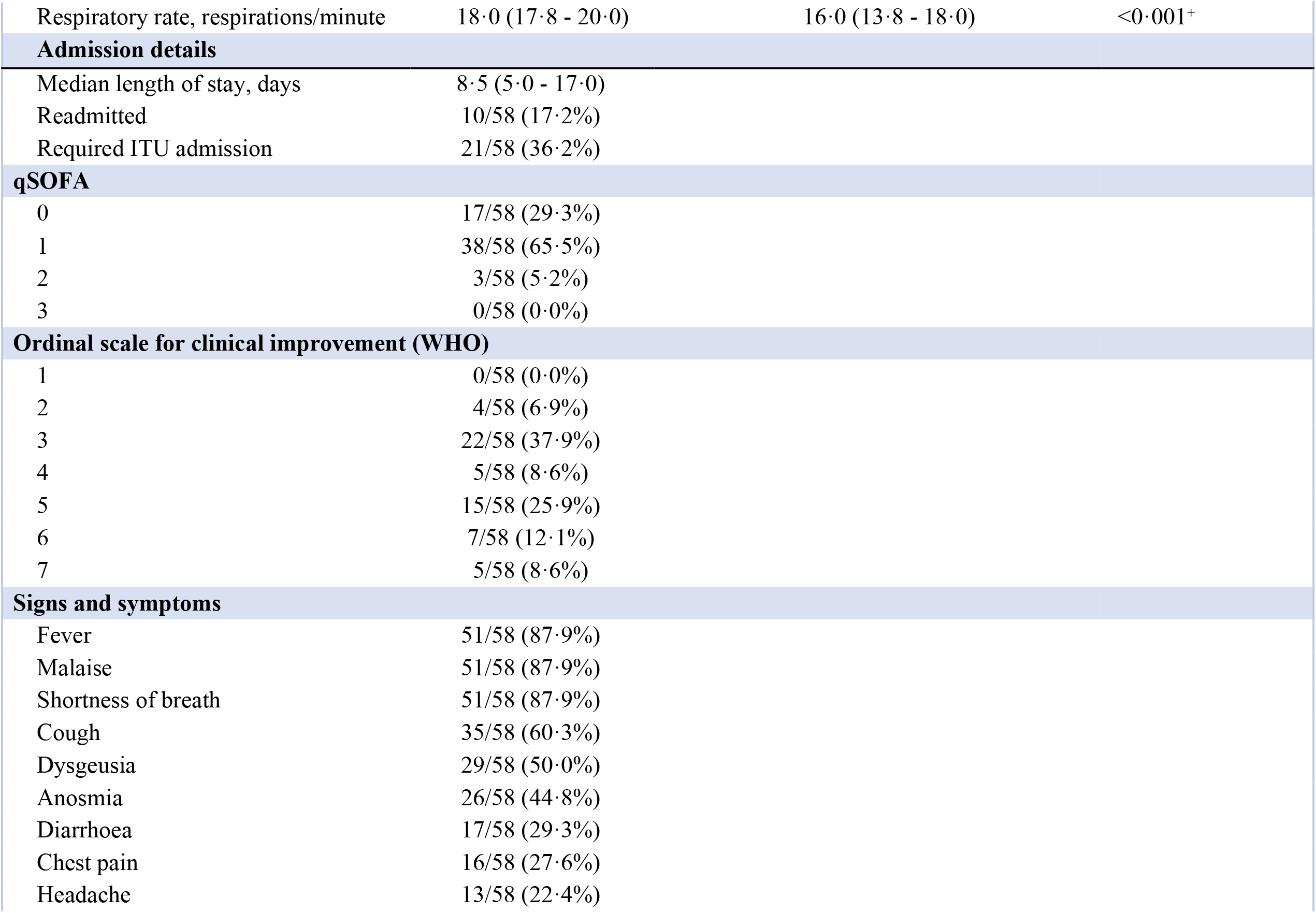
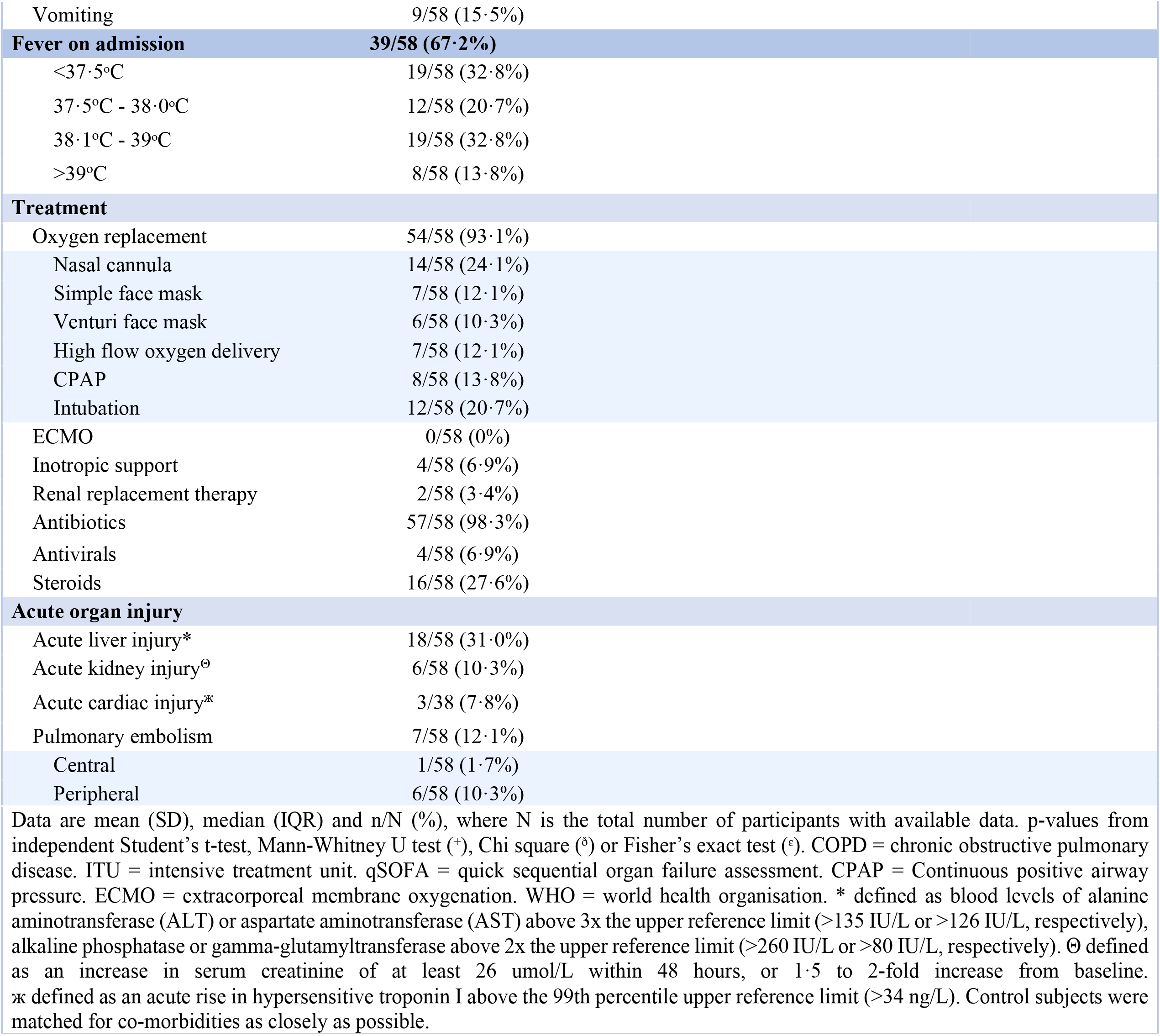
Demographics, baseline characteristics, vital signs at follow-up and admission details of COVID-19 survivors and control participants

|  | COVID | CONTROL | p-value |
| --- | --- | --- | --- |
| **General demographics** |  |  |  |
| Age, years | 55.4 (13.2) | 53.9 (12.3) | 0.62 |
| Sex |  |  | 1.00<sup>d</sup> |
| Female | 24/58 (41.4%) | 12/30 (40.0%) |  |
| Male | 34/58 (58.6%) | 18/30 (60.0%) |  |
| BMI, kg/m<sup>2</sup> | 30.8 (26.2 - 36.4) | 27.3 (23.1 - 35.1) | 0.17<sup>+</sup> |
| Black/Asian and minority ethnic groups | 13/58 (22.4%) | 1/30 (3.3 %) | 0.03 |
| Current/Ex-smoker | 20/58 (34.5%) | 7/30 (23.3%) | 0.34<sup>e</sup> |
| Type 1 Diabetes | 1/58 (1.7%) | 0/30 (0.0%) | 1.00<sup>e</sup> |
| Type 2 Diabetes | 8/58 (13.8%) | 3/30 (10.0%) | 0.74<sup>e</sup> |
| Hypertension | 22/58 (37.9%) | 9/30 (30.0%) | 0.49<sup>e</sup> |
| Coronary artery disease | 2/58 (3.4%) | 0/30 (0.0%) | 0.55<sup>e</sup> |
| Cerebrovascular Disease | 1/58 (1.7%) | 0/30 (0.0%) | 1.00<sup>e</sup> |
| Asthma | 20/58 (34.5%) | 6/30 (20.0%) | 0.22<sup>e</sup> |
| COPD | 3/58 (5.2%) | 0/30 (0.0%) | 0.55<sup>e</sup> |
| Previous cancer | 2/58 (3.4%) | 3/30 (10.0%) | 0.33<sup>e</sup> |
| Depression | 3/58 (5.2%) | 1/30 (3.3%) | 1.00 |
| **Vital signs at follow-up** |  |  |  |
| Heart rate, bpm | 76.3 (14.1) | 70.2 (12.1) | 0.047 |
| Systolic pressure, mmHg | 139.7 (16.5) | 137.2 (17.0) | 0.51 |
| Diastolic pressure, mmHg | 79.5 (71.8 - 86.8) | 71.5 (63.0 - 87.8) | 0.12<sup>+</sup> |
| Temperature, °C | 36.6 (36.5 - 36.7) | 36.5 (36.4 - 36.6) | 0.047<sup>+</sup> |
| Oxygen saturation, % | 96.0 (95.0 - 97.0) | 97.0 (96.0 - 98.0) | 0.008<sup>+</sup> |
| Respiratory rate, respirations/minute | 18·0 (17·8 - 20·0) | 16·0 (13·8 - 18·0) | <0·001<sup>+</sup> |
| **Admission details** |  |  |  |
| Median length of stay, days | 8·5 (5·0 - 17·0) |  |  |
| Readmitted | 10/58 (17·2%) |  |  |
| Required ITU admission | 21/58 (36·2%) |  |  |
| **qSOFA** |  |  |  |
| 0 | 17/58 (29·3%) |  |  |
| 1 | 38/58 (65·5%) |  |  |
| 2 | 3/58 (5·2%) |  |  |
| 3 | 0/58 (0·0%) |  |  |
| **Ordinal scale for clinical improvement (WHO)** |  |  |  |
| 1 | 0/58 (0·0%) |  |  |
| 2 | 4/58 (6·9%) |  |  |
| 3 | 22/58 (37·9%) |  |  |
| 4 | 5/58 (8·6%) |  |  |
| 5 | 15/58 (25·9%) |  |  |
| 6 | 7/58 (12·1%) |  |  |
| 7 | 5/58 (8·6%) |  |  |
| **Signs and symptoms** |  |  |  |
| Fever | 51/58 (87·9%) |  |  |
| Malaise | 51/58 (87·9%) |  |  |
| Shortness of breath | 51/58 (87·9%) |  |  |
| Cough | 35/58 (60·3%) |  |  |
| Dysgeusia | 29/58 (50·0%) |  |  |
| Anosmia | 26/58 (44·8%) |  |  |
| Diarrhoea | 17/58 (29·3%) |  |  |
| Chest pain | 16/58 (27·6%) |  |  |
| Headache | 13/58 (22·4%) |  |  |

|  |  |
| --- | --- |
| Vomiting | 9/58 (15.5%) |
| **Fever on admission** | **39/58 (67.2%)** |
| <37.5°C | 19/58 (32.8%) |
| 37.5°C - 38.0°C | 12/58 (20.7%) |
| 38.1°C - 39°C | 19/58 (32.8%) |
| >39°C | 8/58 (13.8%) |
| **Treatment** |  |
| Oxygen replacement | 54/58 (93.1%) |
| Nasal cannula | 14/58 (24.1%) |
| Simple face mask | 7/58 (12.1%) |
| Venturi face mask | 6/58 (10.3%) |
| High flow oxygen delivery | 7/58 (12.1%) |
| CPAP | 8/58 (13.8%) |
| Intubation | 12/58 (20.7%) |
| ECMO | 0/58 (0%) |
| Inotropic support | 4/58 (6.9%) |
| Renal replacement therapy | 2/58 (3.4%) |
| Antibiotics | 57/58 (98.3%) |
| Antivirals | 4/58 (6.9%) |
| Steroids | 16/58 (27.6%) |
| **Acute organ injury** |  |
| Acute liver injury* | 18/58 (31.0%) |
| Acute kidney injury<sup>⊙</sup> | 6/58 (10.3%) |
| Acute cardiac injury<sup>✱</sup> | 3/38 (7.8%) |
| Pulmonary embolism | 7/58 (12.1%) |
| Central | 1/58 (1.7%) |
| Peripheral | 6/58 (10.3%) |

Data are mean (SD), median (IQR) and n/N (%), where N is the total number of participants with available data. p-values from independent Student's t-test, Mann-Whitney U test (+), Chi square (°) or Fisher's exact test (°). COPD = chronic obstructive pulmonary disease. ITU = intensive treatment unit. qSOFA = quick sequential organ failure assessment. CPAP = Continuous positive airway pressure. ECMO = extracorporeal membrane oxygenation. WHO = world health organisation. \* defined as blood levels of alanine aminotransferase (ALT) or aspartate aminotransferase (AST) above 3x the upper reference limit (>135 IU/L or >126 IU/L, respectively), alkaline phosphatase or gamma-glutamyltransferase above 2x the upper reference limit (>260 IU/L or >80 IU/L, respectively). Θ defined as an increase in serum creatinine of at least 26 µmol/L within 48 hours, or 1.5 to 2-fold increase from baseline. ж defined as an acute rise in hypersensitive troponin I above the 99th percentile upper reference limit (>34 ng/L). Control subjects were matched for co-morbidities as closely as possible.

MRI data (**Table 2**) were available for up to 54/58 patients [brain MRI (n=54), lung MRI (n=53) and cardiac and abdominal MRI (n=52)] and 28/30 controls (Study flowchart and summary of missing data in **appendix Figure 1, p27**, and **appendix Table 7, p25**).

**Table 2.**
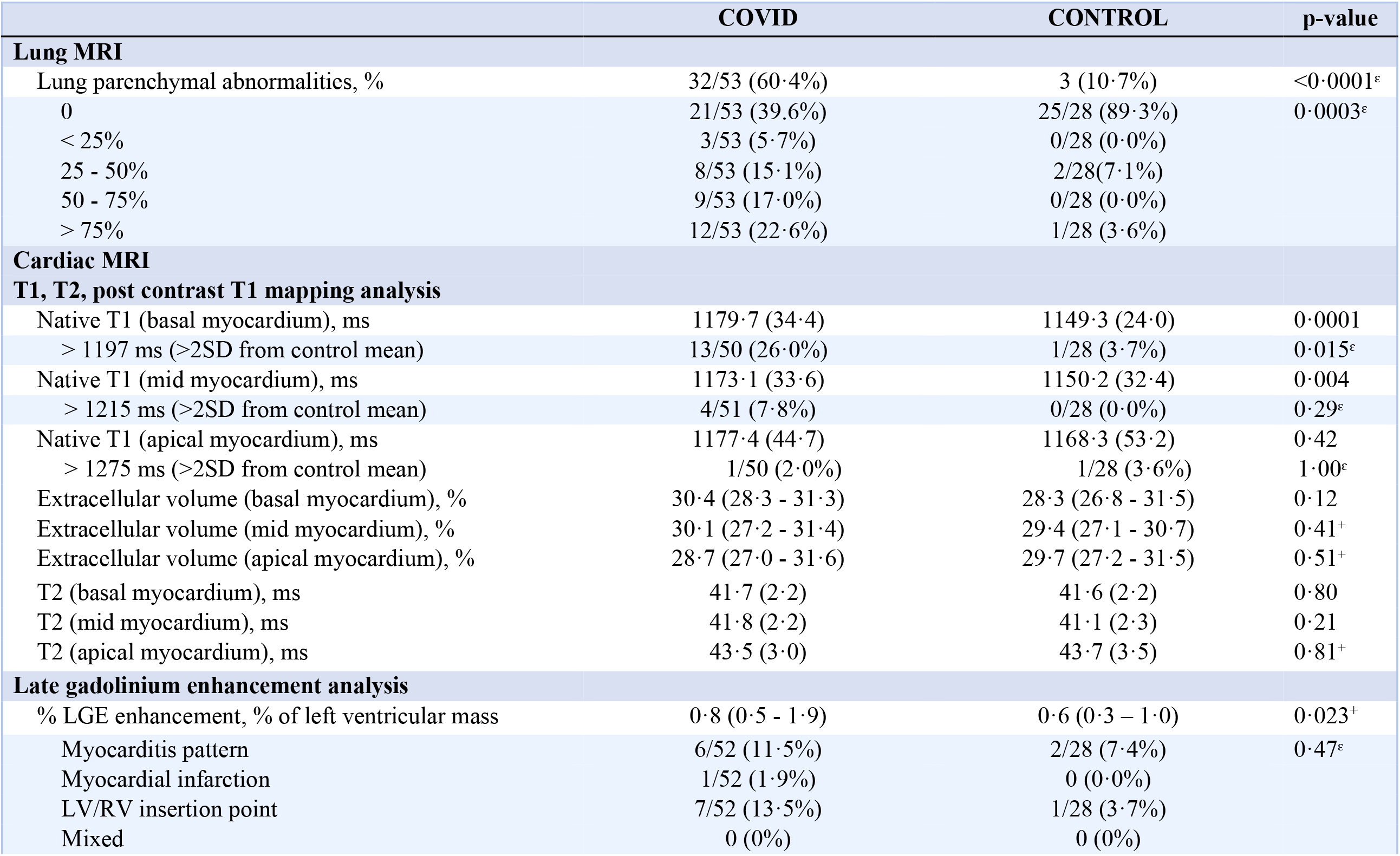
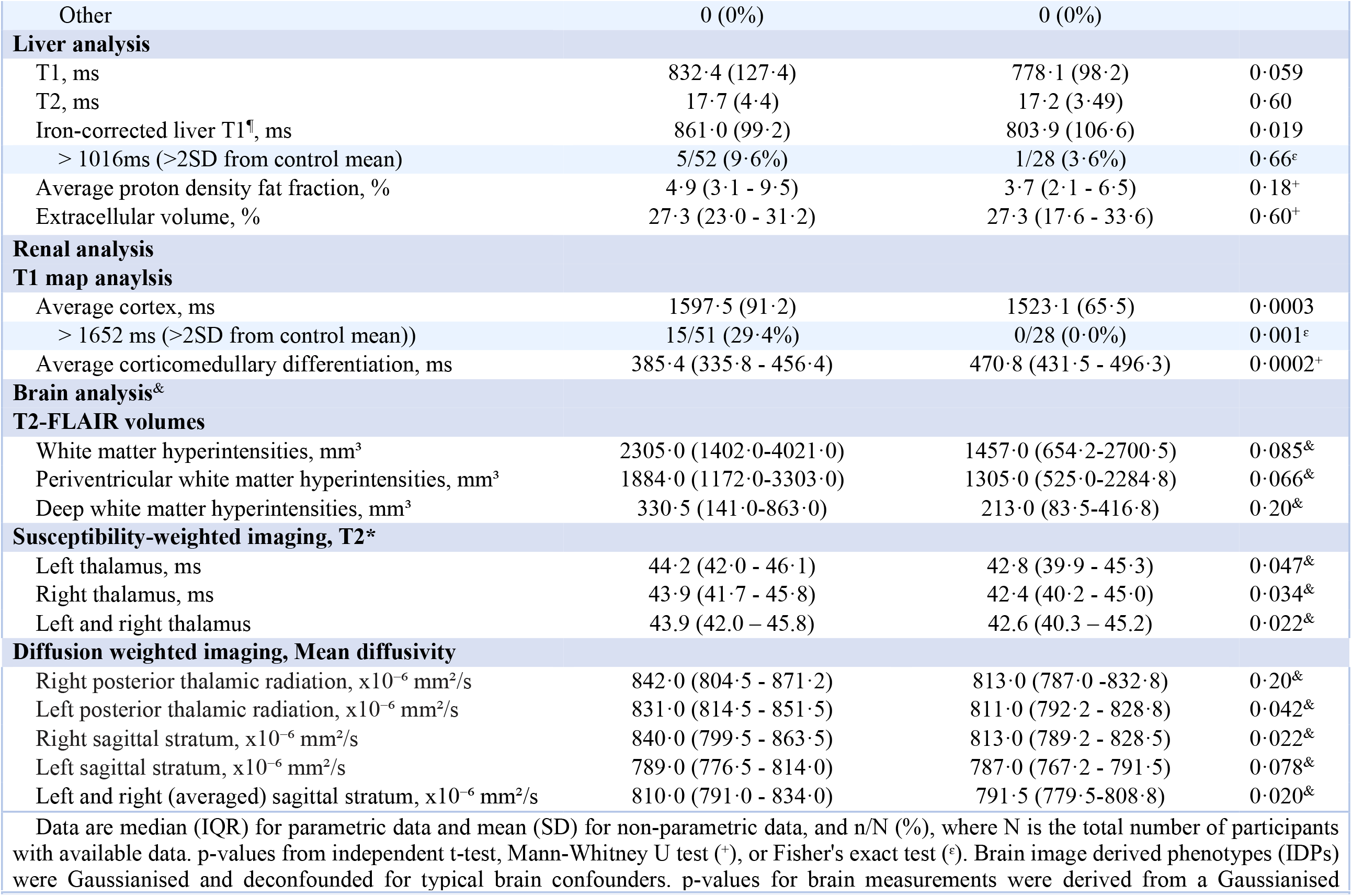
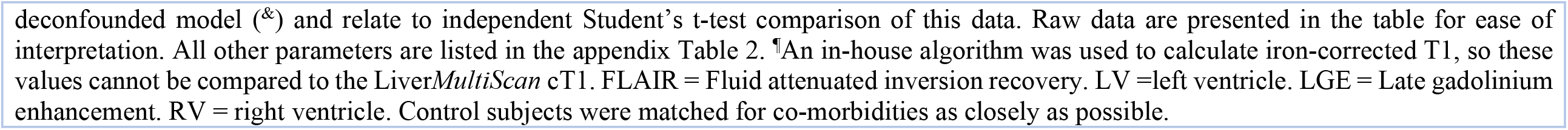
Relevant MRI parameters in COVID-19 survivors and controls

### Lung health and exercise tolerance

During hospital admission, 54/58 (93%) patients had abnormal chest X-ray or computed tomography (CT). At ∼2-3 months, persistent parenchymal abnormalities on lung MRI were present in 32/53 (60%) patients (**Table 2, appendix Figure 2, p28**). Thirty-six (64%) experienced symptoms of significant breathlessness (MRC dyspnoea score ≥2) and 30/55 (55%) complained of fatigue (FSS ≥4). On average, COVID-19 survivors had a significantly lower FEV_1_, FVC and higher FEV_1_/FVC ratio at follow-up (**Table 3**). Abnormalities were noted in FEV_1_ % predicted in 6/56 (11%), FVC % predicted in 7/56 (13%). Patients covered a shorter distance on 6MWT than controls (405±118m vs 517±106m, p<0.0001). Four (7%) patients desaturated at the end of the test. During CPET, patients achieved lower peak maximal oxygen uptake (VO_2_) (**Figure 1**), % of max VO_2_ predicted, anaerobic threshold (% of predicted VO_2_ max), oxygen uptake efficiency slope and higher ventilatory equivalent for carbon dioxide (VE/VCO_2_ slope) (**Table 3, Figure 1**) compared to controls. VE/VCO_2_ slope, a marker of ventilatory efficiency, was worse in those with MRI lung parenchymal abnormalities versus those without (Median 35, IQR (32 - 43) versus 32, IQR (29 - 34), p=0·007). CPET was stopped early in 15/51 (29%) patients due to fatigue and myalgia and 5/51 (10%) of patients due to breathlessness. Anaerobic threshold was reached ealier in patients. VE/VCO_2_ and six-minute walk distance correlated with markers of inflammation in COVID-19 patients (**Figure 1, appendix Table 4, p21**).

**Table 3.** Spirometry and cardiopulmonary exercise test results from patients and controls

| **Spirometry** | **COVID** | **CONTROL** | **p-value** |
| --- | --- | --- | --- |
| FVC, % predicted | 108.3 (22.8) | 131.4 (21.8) | <0.0001 |
| < 80% | 7/56 (12.5%) | 0/28 | 0.090<sup>ε</sup> |
| FEV<sub>1</sub>, % predicted | 101.4 (19.7) | 118.7 (22.1) | 0.0004 |
| < 80% | 6/56 (10.7%) | 1/28 (3.6%) | 0.42<sup>ε</sup> |
| FEV<sub>1</sub>/FVC | 0.77 (0.73 - 0.80) | 0.75 (0.70 - 0.78) | 0.027<sup>+</sup> |
| FEF<sub>25</sub>, % predicted | 97.0 (27.6) | 110.1 (30.4) | 0.020 |
| FEF<sub>50</sub>, % predicted | 81.0 (23.2) | 86.9 (24.5) | 0.13 |
| FEF<sub>75</sub>, % predicted | 54.5 (42.8 - 70.0) | 54.0 (48.5 - 69.5) | 0.60<sup>+</sup> |
| Peak expiratory flow, % predicted | 105.7 (27.7) | 114.5 (24.7) | 0.16 |
| **Cardiopulmonary exercise test** |  |  |  |
| VO<sub>2</sub> peak, % of predicted VO<sub>2</sub> max | 80.5 (23.1) | 112.7 (27.0) | <0.0001 |
| < 80% | 28/51 (54.9%) | 2/27 (7.4%) | <0.0001<sup>ε</sup> |
| Anaerobic threshold (% of predicted VO<sub>2</sub> max) | 40.7 (36.2 - 47.5) | 46.8 (43.3 - 51.3) | 0.0005<sup>+</sup> |
| VE/VCO<sub>2</sub> Slope | 33.4 (29.2 - 40.3) | 28.2 (26.7 - 30.0) | <0.0001<sup>+</sup> |
| Oxygen Uptake Efficiency Slope | 1.9 (1.6 - 2.4) | 2.7 (2.0 - 3.2) | 0.001<sup>+</sup> |
| Data are median (IQR) for parametric data and mean (SD) for non-parametric data, and n/N (%), where N is the total number of participants with available data. p-values from independent Student's t-test, Mann-Whitney U test (+), or Fisher's exact test (ε). FVC = Forced vital capacity. FEV<sub>1</sub> = Forced expiratory volume in 1 second. FEF<sub>25</sub>, FEF<sub>50</sub>, FEF<sub>75</sub> = Forced expiratory flow at 25%, 50% and 75% of forced expiration, respectively. VO<sub>2</sub> = oxygen consumption. VE/VCO<sub>2</sub> = ventilatory equivalent for carbon dioxide. Control subjects were matched for co-morbidities as closely as possible. |  |  |  |

**Figure 1.**
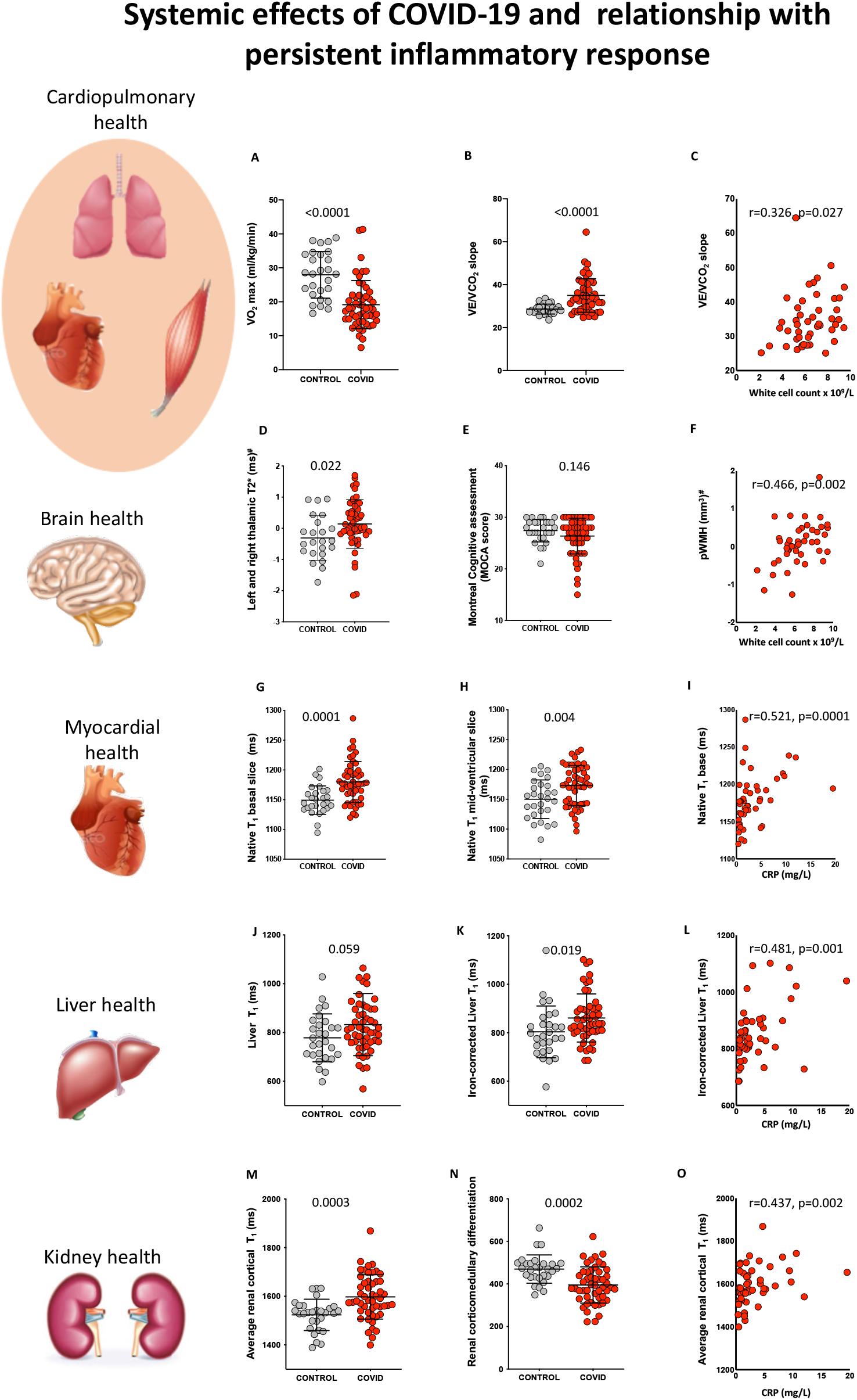
Systemic effects of COVID-19 and relationship with inflammatory response. A, B: Comparison of cardiopulmonary exercise test (CPET) parameters (VO_2_ max and VE/VCO_2_) between comorbidity-matched control and COVID-19 survivors. C: Relationship between VE/VCO_2_ and white cell count in COVID-19. D, E: Comparison of susceptibility weighted T2* signal (left and right thalamus) and MoCA scores between control and COVID-19 survivors. F: Relationship between periventricular white matter hyperintensity volume (pWMH’s) volume and white cell count in COVID-19. G, H: Comparison of myocardial native T1 (base and mid ventricle) between control and COVID-19 survivors. I: Relationship between basal native T_1_ and C-reactive protein (CRP). J, K: Comparison of liver T_1_ and iron-corrected liver T1 between control and COVID-19 survivors. An in-house algorithm was used to calculate iron-corrected T1, so these values cannot be compared to the Liver*MultiScan* cT1. L: Relationship between iron-corrected liver T_1_ and CRP in COVID-19. M, N: Comparison of average cortical kidney T_1_ and corticomedullary differentiation in control and COVID-19 survivors. O: Relationship between average cortical kidney T_1_ and CRP in COVID-19 (p-values for comparisons are from Student’s t-tests for all variables; Spearman’s correlation coefficient and p-values are reported for correlations, # signifies p-values were derived from comparison of variables that were Gaussianised and deconfounded).

### Brain health and cognition

During hospital admission, one patient developed a right occipital stroke; follow-up brain MRI revealed signs of a mature infarct. Blinded qualitative assessment (by expert neuroradiologist) of brain MRI at 2-3 months did not reveal group differences in burden of small vessel disease, white matter hyperintensities, haemorrhage or ischaemic changes between patients and controls (**appendix Table 3, p20**). Quantitative measurements of grey matter volumes (globally and regionally), white matter volumes and cerebral perfusion were not different between groups (**appendix Table 2, p12**). Compared to controls, COVID-19 patients had a higher T_2_* signal on susceptibility-weighted imaging in the left and right thalamus (**Figure 1**) and increased mean diffusivity in the left posterior thalamic radiation and sagittal stratum (left and right averaged). Patients also showed a trend for higher quantitative measure of white matter hyperintensity volume on T_2_-FLAIR imaging compared to controls (**Table 2**).

Cognitive performance in the visuospatial/executive domain was impaired among patients compared to controls (MoCA visuospatial score ≤4 in 40% patients versus 16% in controls, p=0·01). Among patients, 28% (16/58) had a total MoCA score that was abnormal according to the established cut-off of <26 compared to 17% (5/30) of controls. Median MoCA scores in patients (27, IQR 25 - 29), however, were not statistically significantly different from controls (28, IQR 27 - 29, p=0·146) (**Figure 1**). Periventricular white matter hyperintensities and right thalamic T_2_* correlated with markers of inflammation in patients (**appendix Table 4, p21**), but not with cognitive performance.

### Cardiac health

During admission, 38/58 (66%) were screened for cardiac involvement with troponin (high sensitivity Troponin I). Three (8%) were found to have an elevated troponin during admission. At follow-up, troponin was normal in all patients (**appendix Table 1, p9)**. Left ventricular function was normal and comparable between groups. Right ventricular ejection fraction in patients ranged from 43 to 79%, and on average was normal and not different from controls (**appendix Table 2, p12**). Slice-averaged basal and mid-ventricular native T_1_, a marker of fibrosis or inflammation on cardiac MRI, was significantly elevated in patients (**Table 2, Figure 1**). Basal myocardial T_1_ was elevated (> 2 SD from average control T_1_) in 26% (13/50) of patients. Mid myocardial T1 was elevated in 8% (4/51) and average of base and mid myocardial T1 in 24% (12/50) of patients. Native T2, a marker of oedema, was not different between patients and controls. Extracellular volume fraction (a measure of diffuse fibrosis) tended to be higher in the base but the numerical differences failed to reach statistical significance (**Table 2**). Focal fibrosis burden was mildly increased in patients. Both basal and mid-ventricular myocardial T1 correlated moderately (**appendix, p2** for definition) with CRP and pro-calcitonin in patients (**appendix Table 4, p21, Figure 1**), but not in controls (p>0·1).

### Liver health

Acute liver injury (**appendix, p2**) was seen in 31% (18/58) of patients. At 2-3 months, 11% had persistent liver injury (non-specific pattern) on blood tests (**appendix Table 1, p9**). On MRI, another 10% (5/52) of patients had signs of liver injury evidenced by increased iron-corrected liver T_1_ (**Table 2, Figure 1**). Iron-corrected liver T_1_ is a histologically validated imaging biomarker of hepatic fibro-inflammation,^15^ which has subsequently been developed in Liver*MultiScan* where it is increasingly being used to monitor the response of hepatitis to novel therapies.^16^ By contrast, liver fat, iron and extracellular volume fraction (a marker of diffuse fibrosis) did not differ between groups. Iron-corrected liver T_1_ correlated moderately with systemic markers of inflammation (white cell count, neutrophil, monocyte count and CRP) in patients (**appendix Table 4, p21**), but also in controls.

### Haematological system and spleen

Haematological abnormalities including lymphocytopenia and thrombocytopenia were seen in 47% (27/58) and 2% (1/58) of patients at admission respectively and an elevated CRP (>10mg/L) was seen in 98% (57/58). At follow-up, all abnormalities in lymphocyte and platelet count returned to normal. However, patients tended to have higher a CRP (p=0·058) (**appendix, Table 1, p9**). Splenic volume and tissue characteristics on MRI did not differ significantly between patients and controls (**appendix Table 2, p12**).

### Kidney health

Six (10%) patients developed acute kidney injury (**Table 1**), of whom two required renal replacement therapy during admission. At 2-3 months, 3% (2/58) of patients had residual renal impairment which was not present prior to COVID-19. On average, creatinine and estimated glomerular filtration rate were not significantly different between patients and controls (**appendix Table 1, p9**). Despite this, both average (left and right) renal cortical T_1_ and corticomedullary differentiation, markers of renal injury/fibro-inflammation, were abnormal in patients (**Table 2, Figure 1**). A significantly higher renal cortical T_1_ (> 2 SD than control mean) was seen in 29% (15/51) of patients. Patients with acute kidney injury during admission had a higher average renal cortical T_1_ (1711±90ms versus 1582±81ms, p=0·001) and lower corticomedullary differentiation (318±59 versus 405±83, p=0·016; **appendix Figure 2, p28)** compared to those without. Kidney oxygenation did not differ between patients and controls (**appendix Table 2, p12**). Average renal cortical T_1_ had a moderate correlation with markers of inflammation (CRP, pro-calcitonin) in patients at follow-up (**appendix Table 4, p21**).

### Mental health and quality of life

At 2-3 months, a higher proportion of COVID-19 patients reported moderate to severe symptoms of depression (i.e., PHQ-9 score ≥6, 39% versus 17%, p=0·036) and anxiety (i.e, GAD-7 score ≥6, 35% versus 10%, p=0·012) compared to controls (**Table 4, Figure 2**). Patients reported a significantly reduced quality of life in all domains (**Table 4, Figure 2**). Importantly, impairment in both physical and emotional health imposed significant role limitations among COVID-19 survivors. The severity of depression and anxiety did not consistently associate with markers of inflammation (except for monocyte count) or multiorgan injury among COVID-19 patients (**appendix Table 4, p21**). However, a moderate correlation was seen between extent of mood symptoms and anxiety and ongoing breathlessness (MRC dyspnoea score) (PHQ-9: r=0·58, p<0·0001, GAD-7: r=0·41, p=0·002).

**Table 4.** Anxiety (GAD-7), depression (PHQ-9), quality of life (SF-36) and symptom (dyspnoea, fatigue) burden in patients and controls.

|  | COVID | CONTROL | p-value |
| --- | --- | --- | --- |
| **GAD-7** |  |  |  |
| Score | 2.0 (0.0 - 7.5) | 0.5 (0.0 - 4.3) | 0.066<sup>+</sup> |
| 0 - 5 (None - Mild) | 37/57 (64.9%) | 27/30 (90%) |  |
| 6 - 10 (Moderate) | 13/57 (22.8%) | 2/30 (6.7%) |  |
| 11 - 15 (Moderate - Severe) | 4/57 (7.0%) | 1/30 (3.3%) |  |
| ≥16 (Severe) | 3/57 (5.3%) | 0/30 (0.0%) |  |
| Moderate or worse anxiety |  |  |  |
| <6 (None - Mild) | 37/57 (64.9%) | 27/30 (90.0%) | 0.012<sup>ε</sup> |
| ≥6 (Moderate or more) | 20/57 (35.1%) | 3/30 (10.0%) |  |
| **PHQ-9** |  |  |  |
| Score | 3.0 (1.0 - 7.5) | 1.5 (0.0 - 5.0) | 0.009<sup>+</sup> |
| 0 - 5 (None - Mild) | 35/57 (61.4%) | 25/30 (83.3%) |  |
| 6 - 10 (Moderate) | 12/57 (21.1%) | 5/30 (16.7%) |  |
| 11 - 15 (Moderate - Severe) | 9/57 (15.8%) | 0/30 (0.0%) |  |
| ≥16 (Severe) | 1/57 (1.8%) | 0/30 (0.0%) |  |
| Moderate or worse mood symptoms |  |  |  |
| <6 (None - Mild) | 35/57 (61.4%) | 25/30 (83.3%) | 0.036<sup>ε</sup> |
| ≥6 (Moderate or more) | 22/57 (38.6%) | 5/30 (16.7%) |  |
| **SF-36 Domains** |  |  |  |
| Physical Functioning | 65.0 (45.0 - 90.0) | 92.5 (83.8 - 100.0) | <0.0001<sup>+</sup> |
| Role Limitations Due to Physical Health | 25.0 (0.0 - 75.0) | 100.0 (100.0 - 100.0) | <0.0001<sup>+</sup> |
| Role Limitations Due to Emotional Health | 33.3 (0.0 - 100.0) | 100.0 (100.0 - 100.0) | <0.0001<sup>+</sup> |
| Energy | 45.0 (25.0 - 70.0) | 65.0 (55.0 - 80.0) | <0.0001<sup>+</sup> |
| Emotional Well-Being | 76.0 (62.0 - 88.0) | 84.0 (72.0 - 92.0) | 0.044<sup>+</sup> |
| Social Functioning | 50.0 (37.5 - 87.5) | 100.0 (62.5 - 100.0) | 0.0002<sup>+</sup> |
| Pain | 67.5 (35.0 - 90.0) | 85.0 (67.5 - 100.0) | 0.003<sup>+</sup> |
| General Health | 68·8 (43·8 - 81·3) | 75·0 (60·9 - 87·5) | 0·022<sup>+</sup> |
| **Dyspnoea - 12 symptom score** |  |  |  |
| Median (IQR) | 4·0 (1·0 - 11·0) | 0·0 (0 - 1·5) | <0·0001<sup>+</sup> |
| **Fatigue Severity Scale** |  |  |  |
| Median (IQR) | 34 (18-49) | 17 (11-24) | 0·001<sup>+</sup> |
| ≥4 | 30/55 (54·5%) | 5/29 (17·2%) | 0·010<sup>ε</sup> |
| **Medical Research Council Scale** |  |  |  |
| MRC grade 2 - 5 | 36/56 (64·3%) | 3/29 (10·3%) | <0·0001<sup>ε</sup> |
| Data are median (IQR) and n/N (%), where N is the total number of participants with available data. p-values from Mann-Whitney U test (<sup>+</sup>) or Fisher's exact test (<sup>ε</sup>). GAD-7 = Generalised anxiety disorder-7 assessment. PHQ-9 = Patient health questionnaire-9 assessment. SF-36 = Short form 36. MRC = Medical Research Council Scale. Control subjects were matched for co-morbidities as closely as possible. |  |  |  |

**Figure 2.**
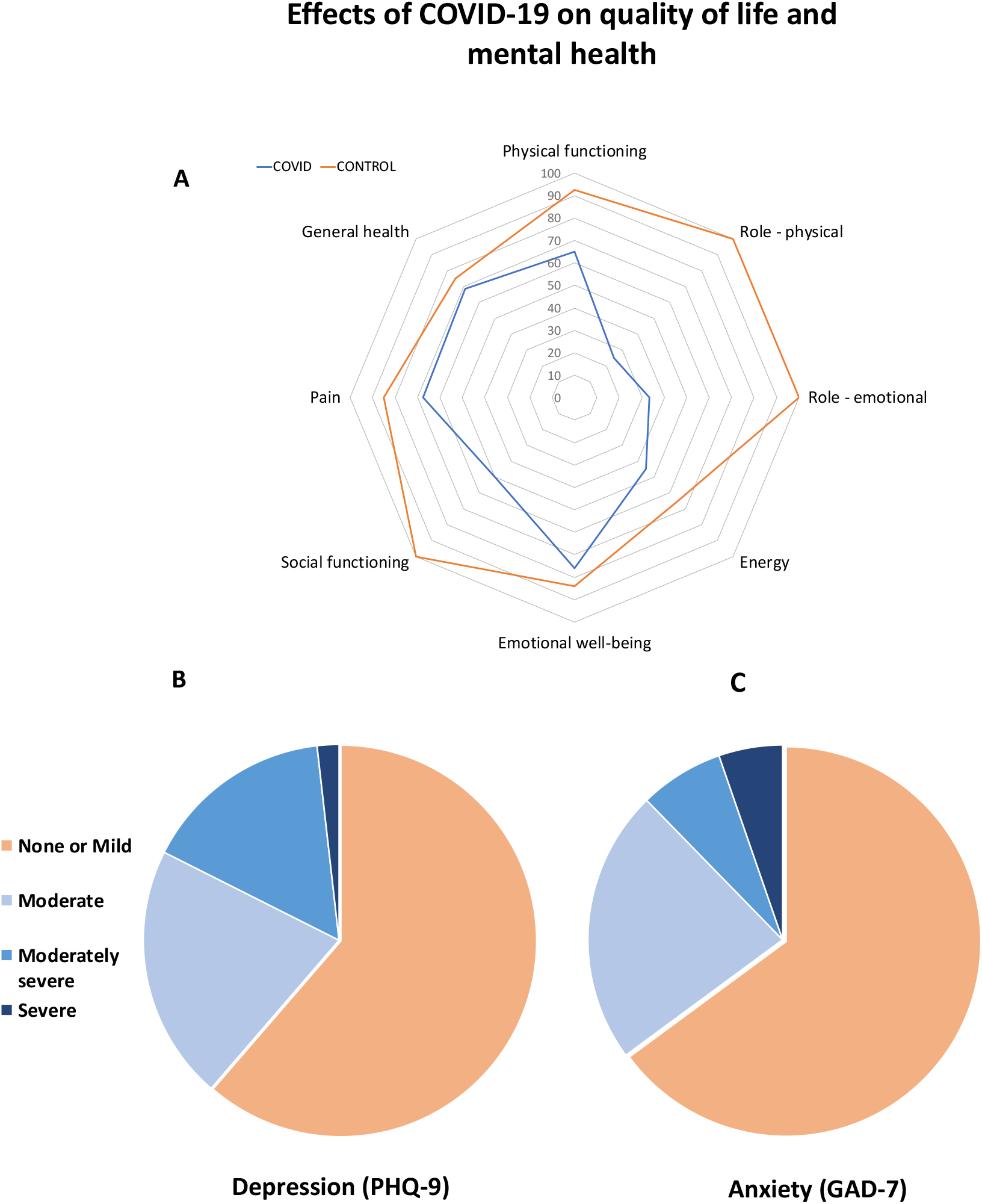
A: Quality of life (Short Form-36) radar plot for COVID-19 survivors and controls. B,C: Burden of depression and anxiety among patients with COVID-19. A: The radar plot demonstrates that patients with COVID-19 (blue line) were more likely to experience impairment in energy, general health, physical health, social and emotional well-being and increased pain when compared to controls (orange line). Both physical and emotional factors caused significant role limitations among patients. B, C: Moderate to severe self-reported symptoms of depression and anxiety were also reported by a third of patients who were hospitalised with COVID-19.

### Severity of disease and persistent inflammation

Severity of illness during admission (WHO ordinal scale) (**appendix Table 4, p21, Table 5, p23**) correlated moderately with inflammatory markers at follow-up, signs of persistent inflammation/injury in the lungs, liver, kidneys, and VE/VCO_2_ on CPET. Notably, even in patients who were not critically ill (i.e, those who were not intubated or ventilated or receiving vasopressor/ionotropic support or renal replacement therapy), MRI evidence of lung, cardiac, kidney and brain abnormalities could be seen (**appendix Table 6, p24)**. Although the severity of illness during admission did not predict the risk of depression, the extent of persistent symptoms did.

## Discussion

The present holistic study uniquely characterised the medium-term effects of COVID-19 infection on multiple vital organs, functional capacity, mental health and cognition in post-hospital survivors of moderate to severe infection. The key findings of our study are: First, at 2-3 months from disease-onset, a proportion of COVID-19 patients displayed abnormalities in the lungs, brain, heart, liver and kidneys on MRI. Second, the severity of acute illness during admission correlated with markers of multiorgan injury at follow-up. Third, limitations in exercise tolerance (CPET and six-minute walk distance) and imaging biomarkers correlated with the extent of persistent inflammation. Deconditioning, symptoms of persistent breathlessness, and fatigue were prominent among patients and interfered with activities of daily living and quality of life. Finally, moderate to severe self-reported depression and anxiety symptoms were more common among patients than controls and were related to the burden of persistent breathlessness at follow-up.

Studies examining the temporal evolution of lung abnormalities on serial high-resolution CT scans have revealed that persistent inflammatory changes may be seen in up to 71% of COVID-19 survivors at three months post discharge.^7,17^ Consistent with this, we observed a high proportion of parenchymal abnormalities on lung MRI, albeit at a lower frequency than that seen on CT. Previous investigations have shown that survivors of SARS pneumonia can be left with more permanent damage to the lungs^18^ and abnormalities in lung function for months to even years after infection. In our study, 13% of patients exhibited abnormalities on spirometry (FVC) at 2-3 months. Although we were unable to assess diffusion capacity (D_LCO_) in our patients, our findings are in line with a recent report by Mo and colleagues, who demonstrated similar anomalies on spirometry and additionally described an impairment in D_LCO_ in up to 47% of cases.^19^

The affinity of SARS-CoV-2 for extra-pulmonary organs is increasingly recognised. Post-mortem studies have confirmed the presence of high titres of SARS-CoV-2 viral load and expression of receptors (Angiotensin-2 Converting Enzymes/ACE2 and Type II transmembrane serine protease/TMPRSS) for viral entry and replication in the lungs, kidneys, liver, heart and brain.^5^ Injury to extrapulmonary organs may be mediated by direct viral toxicity, widespread immunological response and/or thromboinflammation exacerbated by endothelitis.^6^

Occult neurological injury has been suspected in COVID-19 due to a high burden of non-specific neurological symptoms.^20^ Although neurological symptoms were frequent (∼50%) in our unselected cohort, imaging evidence of severe neurologic injury on MRI was rare. Nevertheless, patients demonstrated increased bilateral thalamic T_2_* signal on susceptibility-weighted imaging and increased mean diffusivity in posterior thalamic radiations and sagittal stratum. Susceptibility-weighted imaging is often used to detect blood breakdown products and calcification.^21^ That these abnormalities could reflect a higher burden of microvascular events among COVID-19 survivors is tentatively supported by a trend towards an increased volume of white matter hyperintensities among patients. This would be consistent with the higher frequency of cerebrovascular events reported by others.^20^ While the exact pathophysiology underlying the cerebrovascular disease is yet to be clarified, it is possible that a combination of hypercoagulable state acutely and chronic neuroinflammatory processes, supported by the association of white matter hyperintensity volumes,^22^ T_2_* abnormalities and inflammatory markers, play an important role. The cognitive profile observed (primarily dysexecutive) among patients is also consistent with a vascular pattern, replicating previous reports of dysexecutive syndrome in COVID-19 survivors.^23^ Although our cross-sectional study design limits the extent to which causal associations can be made, our findings suggest a potential link between COVID-19 and future risk of cognitive decline.

Evidence of acute myocardial injury can be seen in up to a third of hospitalised patients with moderate to severe SARS-CoV-2 infection and associates with fatal outcomes.^2,24^ Cardiac MRI has been shown to be particularly useful in providing a diagnosis in patients with suspected cardiac involvement.^25^ In a recent study by Puntmann and colleagues,^26^ cardiac MRI showed evidence of a high burden of inflammation (60% of patients), as seen by elevated native T_1_, T_2_ and some biventricular impairment in convalescing COVID-19 patients, a third of whom required hospitalisation. In our study of hospitalised patients, at 2-3 months, only 26% had a significantly elevated native T_1_. Native T_2_ and cardiac function did not differ from our risk-factor matched cohort consistent with an earlier study by Knight and colleagues^25^. A point worth noting is that differences in prevalence estimates on MRI studies may arise from variations in ‘reference ranges’ (normal versus risk-factors matched controls), methodological differences (sequence parameters, field strength, analysis method), and patient characteristics. Our approach to use a risk-factor matched control group (prospectively enrolled under identical scan conditions) as reference suggests that abnormal tissue characteristics on MRI in 26% of patients could not be explained by the mere presence of comorbidities. Furthermore, myocardial native T_1_ moderately correlated with serum markers of inflammatory response, indicating a possible relationship between the extent of ongoing inflammation and myocardial tissue abnormalities.

Several independent investigations^3,4^ have confirmed a high prevalence of acute liver injury in hospitalised COVID-19 patients. Potential mechanisms include hyperinflammatory syndrome, hypoxia-mediated metabolic derangements, venous thrombosis and drug-induced hepatitis.^27^ Direct infection of cholangiocytes has also been suggested, as SARS-CoV-2 may injure the bile ducts by binding to ACE2 receptors.^28^ We observed that 11% of patients had persistent blood biomarker evidence of liver injury at 2-3 months, and another 10% demonstrated increased iron-corrected liver T_1_, a marker of fibroinflammation.^15^ In our study, the extent of liver injury on MRI moderately correlated with inflammatory markers, supporting the role of inflammation in promoting hepatic damage.

The kidneys are amongst the most common targets of SARS-CoV2, with acute kidney injury reported in 0.5-37% of hospitalised COVID-19 patients.^2-4,29,30^ Direct infection of renal cells may occur via ACE2 receptors which are enriched in podocytes and endothelial cells.^31^ Associated histopathological abnormalities include prominent lymphocytic endothelitis, acute tubular necrosis, diffuse erythrocyte aggregation, peritubular obstruction and podocyte injury.^31^ We showed that 29% of patients had abnormal renal tissue characteristics on MRI. In particular, cortical T_1_, a marker of renal inflammation/injury was prolonged and accompanied by a loss of corticomedullary differentiation, a pattern reminiscent of other post-inflammatory glomerulonephritides.^32^ Patients with more severe disease (i.e, acute kidney injury, need for higher oxygen support) and higher inflammatory burden were more likely to have abnormal cortical T_1_ and corticomedullary differentiation at follow-up.

Chronic inflammation represents a sustained reaction of the immune system to an inflammatory stimulus (i.e, viral nucleic acid) accompanied by tissue damage.^33^ The association of multiple imaging markers of organ abnormalities and inflammatory response in COVID-19 patients, but not in controls, suggests that chronic inflammation may play a role in mediating multiorgan abnormalities.^34^ Although critical illness has also been shown in prior studies to associate with systemic inflammation^35^, we found that MRI evidence of multiorgan abnormalities was not limited to patients with critical disease alone. Further efforts to understand the role of specific immunopathological mechanisms underlying this inflammatory process, and strategies to arrest them, could be important in limiting the long-term detrimental effects of COVID-19 on vital organs.

Insights from earlier studies of SARS survivors^36^ have raised concerns that limitations in exercise tolerance may persist for months after infection. In our study, patients achieved a shorter six-minute walk distance, lower peak VO_2_ and lower % of predicted VO_2_ max at the anaerobic threshold (VT1). VE/VCO_2_ slope, a measure of ventilatory efficiency, was worse in patients with parenchymal abnormalities and both VE/VCO_2_ and six-minute walk distance correlated with markers of systemic inflammation. Of note, a higher proportion of patients (29%) stopped the CPET early because of generalised muscle ache and fatigue rather than breathlessness. These findings suggest that muscle wasting, secondary to a catabolic state induced by severe illness^37^ and potentially ongoing inflammation^38^, could be an important contributor to exercise limitations, along with lung parenchymal abnormalities.

In addition to coping with the debilitating acute effects of COVID-19, survivors experience a range of mental stressors whilst in-hospital and after discharge. Not surprisingly, we and others^39,40^ have observed a high-level of self-reported symptoms of anxiety and depression among survivors. Infection-triggered cytokine dysregulation and the neurotropic potential of SARS-CoV-2 have widely been speculated to induce psychopathological sequelae among COVID-19 patients, consistent with neuroinflammatory mechanisms implicated in other psychiatric disorders.^41^ Here, although the burden of ongoing symptoms of breathlessness associated with mood and anxiety symptoms, we did not see a consistent association between markers of systemic inflammation or organ injury and depression. Given the limited sample size of our study, a more focussed approach in a larger cohort could yield further insights into such relationships and offers the potential to identify novel targets for neuropsychiatric therapeutic modulation.

### Limitations

The relatively small sample size of this single-centre study, its cross-sectional design and lack of correction for multiple comparisons are important limitations which curtail the generalisability of our findings and accuracy of prevalence estimates. The lack of pre-COVID imaging also limits our ability to make causal inferences about the mechanism of multiorgan abnormalities in patients recovering from COVID-19 infection. However, this is the first exploratory study to comprehensively undertake a holistic assessment of multiple vital organs, mental, cognitive and physical health in patients with COVID-19 post-hospital discharge. These findings underscore the need for further large scale investigations as is currently planned by Public Health England through the Post-HOSPitalisation COVID-19 (PHOSP-COVID)^42^ national consortium and its two MRI sub-studies C-MORE and COVERSCAN. The assessment of lung function and lung injury were limited given the lack of *D*_LCO_ assessments and CT chest in all subjects, due to restricted access to lung function testing. The use of lung MRI in place of CT could have underestimated the prevalence of lung injury. Controls in our study were not hospitalised, thus group differences in symptomatology, mental health, exercise capacity and quality of life may not be specific to COVID-19. We excluded patients with severe comorbidities to provide more reliable estimates of COVID-19 specific injury and may have underestimated the extent of multiorgan damage in the wider population. There were ethnoracial differences between the control and COVID-19 patients enrolled in this study which may have played a role in prevalence estimates of multiorgan injury. Finally, whether the findings on MRI have any long term clinical implications remains to be determined by further longitudinal follow-up studies.

## Conclusions

Among post-hospital survivors of moderate to severe COVID-19 infection, abnormalities on multiorgan MRI were seen in a proportion of patients at 2-3 months from disease onset. The severity of acute illness and serum markers of inflammation correlated with the extent of multiorgan changes on MRI and reduced exercise tolerance at follow-up. COVID-19 survivors experience a high burden of depression, anxiety, as well as reduced quality of life post-hospital discharge. Our findings underscore the need to further understand the pathophysiological mechanisms underpinning multiorgan MRI tissue abnormalities and to provide a holistic-integrated multidisciplinary model of clinical care for COVID-19 patients post hospital discharge.

## Data Availability

Individual de-identified participant data can be made available when the trial is complete, or upon requests directed to the corresponding author.

## Acknowledgements

The authors’ work was supported by NIHR Oxford and Oxford Health Biomedical Research Centre, Oxford British Heart Foundation (BHF) Centre of Research Excellence, United Kingdom Research Innovation and Wellcome Trust. P.J. thanks the Dunhill Medical Trust for support. The views expressed are those of the authors and not necessarily those of the National Health Service, NIHR, or the United Kingdom Department of Health.

This project is part of a tier 3 study (C-MORE) within the collaborative research programme entitled PHOSP-COVID Post-hospitalisation COVID-19 study: a national consortium to understand and improve long-term health outcomes. Funded by the Medical Research Council and Department of Health and Social Care/National Institute for Health Research Grant (MR/V027859/1) ISRCTN number 10980107.

This work also arises from one of the national “COVID-19 Cardiovascular Disease Flagship Projects” designated by the NIHR-BHF Cardiovascular Partnership.

We thank our patients and their families who have sought to help others understand about the effects of COVID-19. We are grateful to the University of Oxford and Oxford University Hospital Trust for their support of this study. We would like to acknowledge OCMR staff, Ms Polly Whitworth, Ms Joanna Leal, Ms Claudia Nunes, Ms Harriet Nixon, Ms Injung Jang, Ms Maryam Khan, Ms Gillian Anderson, Mr Mike Mcdonald, Ms Jasmine Taylor, Dr Ahmed Moola, Ms Sarah White, Dr Innes Abdel Assam, for their help with this work. We acknowledge the support of Siemens in providing a licence for the use of ASL.

## Data sharing statement

Individual de-identified participant data will be made available when the trial is complete, or upon requests directed to the corresponding author; after approval of a proposal, data can be shared through a secure online platform.

## Author contribution

BR and SN had the idea for and designed the study with the help of all co-authors and had full access to all the data in the study and take responsibility for the integrity of the data and the accuracy of the data analysis. BR and HL were involved in application of ethics. MM, BR, CK, RM and MC collected the data. PJ, EMT, RM, KM, SS, SM, CM were involved in developing the MRI protocol. EOO, MC, LG, SM, FA, TO, FKM, CW, CA, FL, JA, MJ, SS, EMT, XC, BR, AL, FEM performed the analysis. BR, MC and SN drafted the paper. All authors critically revised the manuscript for important intellectual content and gave final approval for the version to be published. All authors agree to be accountable for all aspects of the work in ensuring that questions related to the accuracy or integrity of any part of the work are appropriately investigated and resolved.

