## Appendix for "Medium-term effects of SARS-CoV-2 infection on multiple vital organs, exercise capacity, cognition, quality of life and mental health, post-hospital discharge"

SUPPLEMENTARY APPENDIX

### Methods

#### Inclusion Criteria

We included all patient with moderate to severe (see below for definition) sudden acute respiratory distress syndrome Coronavirus-2 (SARS-CoV-2 infection), confirmed by a reverse transcription and polymerase chain reaction (RT-PCR) nasopharyngeal swab test. Only patients with positive RT-PCR were included in the study.

Controls were non-hospitalised subjects (invited from the community) without symptoms or signs of a respiratory tract infection of coronavirus disease, who were screened for SARS-CoV-2 antibodies and tested negative.

#### Exclusion Criteria

Subjects with contraindications to magnetic resonance imaging (metal implant in body, known claustrophobia, pacemakers, contrast allergy) and severe comorbidities – end-stage renal, cardiac, liver, neurological disease were excluded from the study.

#### Definitions

**Moderate to severe SARS-Co-V2 infection:** Severity of acute illness on admission was defined by the World Health Organisation (WHO) interim clinical guidance. All patients with clinical signs of pneumonia such as respiratory rate > 30 breaths/min; or severe respiratory distress; or SpO2 < 90% (on room air) and requiring hospital admission for more than 48 hours were assessed to have moderate to severe coronavirus disease (COVID-19).[1]

**Severity of disease (or clinical response) during admission:** Patient response in hospital was defined by the WHO ordinal scale for clinical improvement.[2] A score 0 was given to an uninfected individual, 1 to an ambulatory patient without limitations of activities, 2 where there was limitations of activities, 3 for a hospitalised patient not on oxygen ($O_2$) therapy, 4 for $O_2$ supplementation via simple face mask or nasal prongs, 5 for administration of non-invasive ventilation (NIV) or high flow $O_2$, 6 for being intubated and on mechanical ventilation, 7 for requiring ventilation and additional organ support such as renal replacement therapy and extracorporeal membrane oxygenation (ECMO), and 8 for death.[2]

**Lymphocytopenia:** was defined as a lymphocyte count of less than 1000 cells per cubic millimeter.[3]

**Thrombocytopenia:** was defined as a platelet count of less than 100,000 per cubic millimetre.[3]

**Acute kidney injury**: Acute kidney injury (AKI) was defined based on the highest serum creatinine level and urine output. A diagnosis of AKI was made based on any of the following: an increase in serum creatinine levels by 0.3 mg/dl or greater (26.5 μmol/l or greater) within 48 hours; an increase in serum creatinine levels to 1.5 times of the baseline level or greater, which was known or presumed to have occurred within 7 days; or urine volume of below 0.5 ml/kg/h for 6 consecutive hours.[3]

**Acute cardiac injury:** Acute cardiac injury was suspected when serum levels of cardiac biomarkers (high-sensitivity cardiac troponin I) were above the 99$^{th}$ percentile upper limit of normal or new abnormalities were seen on electrocardiography and echocardiography.[3]

**Acute liver injury:** Acute liver injury was defined as blood levels of alanine aminotransferase (ALT) or aspartate aminotransferase (AST) above three times the upper reference limit, alkaline phosphatase or gamma-glutamyl transferase levels above two times the upper reference limit.[4]

**Definition of strength of correlation:** A correlation coefficient of 0 signifies no correlation. Correlation efficient between ±0.1 to (less than) ±0.3 is considered weak.[5] Correlation efficient between ±0.3 to (less than) ±0.7 is considered moderate. A coefficient of ±0.7 to ±0.9 is considered strong and a coefficient of ±1 is perfect.

#### Multiorgan MRI

Scans were carried out during a single scan session on a Siemens Prisma 3T, with a Siemens 20-channel head coil for neuroimaging, and a 32-channel spine array and 30-channel body array used for thoracic and abdominal imaging.

#### Lung MRI

A) Free breathing half-Fourier-acquisition single-shot turbo spin-echo (HASTE) MRI – [Typically axial and coronal; TR/TE = 750/49 ms; R=2; field of view = 380x320 mm,$^2$ slice thickness/spacing 8/0mm, 35 slices, matrix = 256x123]. T2-weighted HASTE images provide high-signal intensity in water-rich tissues. Lung abnormalities tend to appear bright, surrounded by air-filled parenchyma with low signal intensity.

**Lung MRI Analysis**
Axial and coronal thoracic HASTE images were qualitatively read by an expert radiologist (XC) and scored as 0 (0%), 1 (1–25%), 2 (26–50%), 3 (51–75%), or 4 (76–100%) depending on the extent of parenchymal involvement.

**Brain MRI**
The neuroimaging protocol comprised of both structural and functional sequences and lasted approximately 17 minutes. The MRI sequences (brain modalities) were as follows:
A) High-resolution $T_1$-weighted Magnetization Prepared RApid Gradient Echo (MPRAGE) – [TR/TE/TI = 2000/1.95/880 ms, R=2, flip angle = 8°, Matrix = 256x256x208, voxel dimension = 1 mm isotropic]. This sequence is primarily used to study grey matter (GM) structural macroscopic tissue in both cortical and subcortical brain regions. GM reductions have been widely associated with disease and age-related cognitive dysfunction.
B) Diffusion MRI (dMRI) – [TR = 8500 ms, flip angle = 90°, Matrix = 104x104x72, voxel dimension = 2 mm isotropic, b=0, 1000 s/mm$^2$, 3 sequentially applied diffusion gradient directions (x,y,z), blip-reversed b=0]. With a greater number of diffusion-encoding directions, this sequence would measure the random motion of water molecules to allow one to 1) infer information about White Matter (WM) microstructural properties and 2) delineate the gross axonal organisation of the brain. With just 3 orthogonal diffusion directions (as commonly done in clinical practice), we can estimate mean diffusivity. The diffusion sequence was kindly provided by the CMRR, University of Minnesota, USA.
C) Fluid Attenuated Inversion Recovery ($T_2$-FLAIR) – [TR/TE/TI = 5000/386/1800 ms, R=3, Matrix = 256x256x192, voxel dimension = 1x1x1.05 mm]. This sequence is commonly used in clinical practice, for example to characterise white matter (WM) hyperintensities (often referred to as lesions, and arising from a range of biological/pathological mechanisms).
D) Arterial Spin Labelling (ASL) using a multi-slice multi-inversion-time pseudo-continuous ASL (PCASL) sequence[6]– [primaryTR/TE = 4500/14 ms, Matrix = 64x64x24, voxel dimension = 3.4x3.4x4.5 mm, label duration = 1400ms, post-labelling delays: 300/600/900/1200/1500/1800 ms varied in a looped fashion and repeated 5 times, first volume is a calibration ($M_0$) image]. This sequence is used to quantify tissue cerebral blood flow (CBF) and Arterial Transit Time (ATT) by using the water in arterial blood as an endogenous contrast agent.
E) Susceptibility-weighted MR Imaging (3D-swMRI) – [TR/TE1/TE2 = 27/9.4/20 ms, Matrix = 256x232x48, R = 2, voxel dimension = 0.9x0.9x3 mm$^3$]. This sequence is commonly used in clinical settings to study (for example) traumatic brain injury and stroke, but can also be used to define vascular malformations, cerebral microbleeds, intracranial calcifications, neurodegenerative diseases and brain tumours.

**Brain MRI analysis**
All data were manually quality-control checked, resulting in usable brain imaging data from 51 patients and 25 controls. Image processing was largely carried out using a slightly adapted version of the processing pipeline that we have created for the UK Biobank brain imaging.[7-9] This is largely based around tools from FSL (FMRIB Software Library.[10] Data were corrected for gradient and EPI distortions, aligned to each other using linear alignment[11], and then aligned into standard template space (MNI152) using nonlinear alignment applied to the T1.[12,13]
$T_1$ images were further segmented probabilistically into different tissue types[14] and also into different subcortical structures.[15] T2-FLAIR images were also segmented, to identify WM hyperintensities (WMH).[16] pWMH (periventricular WMH) and dWMH (deep WMH) volumes were extracted as subsets of WMH, using the criterion of being less than (or more than, respectively) 10 mm distant from the lateral ventricles.[17,18]

swMRI data was processed in 3 distinct ways: 1) The two echoes were combined to provide a quantitative mapping of $T_2$*. 2) The phase and magnitude data were processed to provide maps highlighting features such as microbleeds. 3) Quantitative susceptibility mapping (QSM) was carried out.[19]
Diffusion data was preprocessed to remove effects of eddy currents, head motion, and slice dropouts,[20] and then processed to generate a mean diffusivity (MD) map.
The ASL data was processed using the BASIL ASL tools in FSL, including modelling of the macrovascular component,[21,22] to estimate CBF maps and AAT. A grey matter mask was defined using a partial volume threshold of 50%; this mask was then used to estimate mean grey matter CBF and AAT.
We estimated a large number of imaging-derived phenotypes (IDPs) - summary measures of brain structure and function [ https://www.fmrib.ox.ac.uk/ukbiobank/ ]. Cross-subject analyses incorporated a range of imaging-related confound covariates: age, sex, BMI, systolic and diastolic blood pressure, smoking status and head size. Areas of the brain with differences in imaging derived phenotypes are highlighted in **appendix Figure 3**.[12]

Qualitative assessment of all brain MRI images was also undertaken by an expert neuroradiologist (FS) who commented on presence of small vessel disease, white matter hyperintensities, cerebral infarcts, bleeds and increased signal in the olfactory cortex (**appendix, Table 3**).

**Cardiac MRI**

The cardiac MRI protocol included routine clinical and advanced parametric mapping sequences. The cardiac MRI sequences were as follows:

A) Cine steady-state free precession (SSFP) imaging - [Three long-axis and short-axis stack; Typically TR/TE = 3.16/1.38ms; R = 3; flip angle = 65°; field of view = 360×270 mm, matrix 208×139, slice thickness/spacing 7/3mm]. This is used in the clinical setting to assess cardiac (left and right ventricular) mass, volumes and function which may be affected in cardiovascular diseases.

B) Shortened Modified Look Locker $T_1$ mapping (ShMOLLI, Siemens prototype sequence WIP1048B) [Base, mid, apex short axis slices; TR/TE = 2.7/1.07ms, R = 2, flip angle = 35°, field of view = 360×270 mm, matrix = 192×144; slice thickness = 8 mm. Inversion times in image acquisitions, waiting periods with conditional on-the-scanner $T_1$ map reconstruction as described previously].[23] This sequence helps assess the myocardial tissue characteristics which indicates if the myocardium has increased water content or fibrosis.[24]

C) $T_2$ mapping (Siemens Myomaps) - [Base, mid, apex short axis; bSSFP readout; TR/TE = 3/1.3ms, R = 2, flip angle = 20°, Matrix = 192×142, field of view = 360×270 mm, slice thickness = 8 mm]. This sequence is typically used to detect increased water content or swelling in myocardial tissue.

D) Late gadolinium imaging was acquired using a $T_1$-weighted phase-sensitive inversion recovery sequence following a bolus injection of 0.15 mmol/kg of body weight of gadolinium-based contrast agent (Dotarem) and a 10 ml saline flush. Typical scan parameters: [TR/TE/TI = 2.9/3.36/subject-specific ms; R = 2; flip angle = 55°; matrix = 144×256, field of view 380×285 mm, slice thickness = 8 mm.] Late gadolinium imaging is used in clinical cardiac MRI to detect regions of focal fibrosis or scar within the myocardium.

E) -$T_1$-weighted imaging: Base, mid and apical $T_1$ maps (ShMOLLI) were acquired 15 minutes after the administration of contrast. This sequence is used to estimate the extent of diffuse fibrosis or scar in the myocardium.

**Cardiac MRI image analysis**

All cardiac MRI images were analysed using cvi42 software (Circle Cardiovascular Imaging Inc., Version 5.10.1, Calgary, Canada).

Volume and function: Left and right ventricular short axis epicardial and endocardial borders were manually contoured at end-diastole and end-systole (endocardial border only). Left and right ventricular end-diastolic (EDV) and end-systolic (ESV) volumes were used to calculate stroke volume (SV) as SV = EDV-ESV.[25] Ejection fraction (EF) was calculated as a ratio of SV and EDV (EF = SV/EDV). LV mass was calculated by subtracting the endocardial volume from the epicardial volume, based on prior knowledge of myocardial specific gravity (1.05 g/cm3).[26] Volumes and mass were indexed to body surface area.

Analysis of $T_1$ and $T_2$ maps: All (pre and post-contrast) T1 maps underwent strict quality control as previously described [24] by observers who underwent standardised institutional training.[27,28] R2 maps of ShMOLLI inversion recovery fit was used to verify the quality of $T_1$-maps on the scanner. Manual epicardial and endocardial contours were drawn on base, mid and apical slices and average slice $T_1$ was derived. Extra-cellular volume (ECV) for the three slices were calculated from pre- and post- contrast $T_1$ maps and haematocrit (HCT) using the formula: $ECV = (\Delta[1/T_1 myocardium]/\Delta[1/T_1 blood]) * [1-Hct])$.

All $T_2$ maps also underwent strict quality control. Manual epicardial and endocardial contours were drawn on base, mid and apical slices (acquired at identical slice positions to pre and post-contrast $T_1$ maps). Average slice $T_2$ was derived for each slice.[29]

Late gadolinium enhancement (LGE): Endocardial and epicardial borders were drawn on the phase sensitive inversion recovery (PSIR) images alone with a reference region of interest (ROI) with normal myocardial intensity as ‘remote’ region. Hyperenhanced pixels were defined as those that were five standard deviations (5-SD) above the mean intensity of the reference ROI placed in a remote area of myocardium with no visual evidence of enhancement.[29]

Blinded qualitative assessment of LGE images were undertaken by an experienced CMR reader (BR) and visually classified as the following: 1) no LGE, 2) myocarditis pattern, 3) myocardial infarction, 4) LV/RV insertion point fibrosis, 5) mixed (infarction and myocarditis), 6) other (cardiomyopathy). All cases were also assessed for presence of pericardial effusion.[30]

**Abdominal MRI**

(A) MOLLI-variant $T_1$ map (Siemens Myomaps) was used to assess fibrosis and inflammation in the liver and spleen – [axial; TR/TE = 2.45/1.05 ms; R=2; flip angle = 35°; field of view 440×330 mm, matrix = 192×144, slice thickness 8mm; ECG-gated shMOLLI sampling pattern and conditional processing]. [23]

(B) Multiecho gradient echo IDEAL[31] was used to assess fat and iron in the liver - [axial; TR/TE = 15/1.1, 2.2, 3.3, 4.4, 5.5, 6.6, 7.7, 8.8, 9.9, 11, 12.1, 13.2 ms; flip angle = 6°; field of view 440×400 mm, matrix = 128×116, slice thickness 10 mm]. These data were reconstructed using the ISMRM fat-water separation toolbox (https://www.ismrm.org/workshops/FatWater12/data.htm) to yield proton density fat fraction (PDFF), $B_0$ and $T_2$* maps.

(C) Kidney MOLLI $T_1$ map with a 5-3s-3 sampling scheme[33] [oblique coronal; TR/TE = 2.7/1.15 ms; R = 4; flip angle = 35°; field of view 320×320 mm, matrix = 194×192, slice thickness 5.5 mm] was used to assess inflammation and fibrosis
(D) Kidney multi echo gradient echo[34] [oblique coronal; TR/TE = 81/4.92, 7.38, 9.84, 12.3, 14.76, 17.22, 19.68, 22.14, 24.6, 27.06, 29.52, 31.98 ms; R=3; flip angle = 25°; field of view 288×288mm, matrix = 192×192, slice thickness 5.0 mm] was used to assess renal oxygenation via R2*.

**Liver and spleen image analysis**
Analysis was performed in the liver by placing three 15 mm diameter circular ROI within the liver parenchyma, as developed by Perspectum,[35] avoiding vessels and bile ducts as much as possible, and any fat/water swaps visible in the PDFF maps. Iron-corrected $T_1$[32] was calculated using our method developed in-house. Control values derived are not from the standardised Liver*MultiScan* device developed by Perspectum and therefore cannot be compared to Liver*MultiScan* cT1. Average values of PDFF, $T_2$*, $T_1$ and iron-corrected $T_1$ for the liver were recorded. A single 15 mm ROI was placed on the spleen, with average $T_1$ and $T_2$* was also recorded. Volumetric segmentation of spleen was carried out using CVI42 (Circle Cardiovascular Imaging, as before). In most subjects, a post-contrast ShMOLLI $T_1$ map was acquired. Circular ROIs of 10 mm diameter were placed on the aorta in the pre- and post-contrast $T_1$ maps, and three 15 mm ROIs in the liver parenchyma as before. The average $T_1$ in these ROIs were used to calculate a liver ECV value, using the same methodology that has been validated in the heart.[36]

**Kidneys image analysis**
$T_1$ mapping was assessed using manually defined ROIs in the cortex and medulla; cortical and medullary $T_1$ were reported along with corticomedullary $T_1$ difference.[37] R2* maps were calculated using a monoexponential fit and the twelve-layer concentric object (TLCO) method[38] used to yield a single renal BOLD parameter, BOLD radial gradient in Hz/%.
**Spirometry**
Spirometry measurements were undertaken in triplicate using a calibrated spirometer (Metalyzer 3B, Cortex Biophysik, Leipzig,Germany), according to the criteria established by the American Thoracic Society and European Respiratory Society.[39] Spirometry measures were not repeated after administration of bronchodilators. The largest $FEV_1$ and FVC of the triplicate readings and respective $FEV_1$/FVC ratio were reported.

**Cardiopulmonary exercise test (CPET)**
All patients performed symptom-limited incremental cycle ergometer exercise testing with electrocardiographic monitoring. The gas analyser was calibrated prior to each test. Each subject was fitted with a facemask incorporating a disposable flow turbine.
Continuous 12-lead electrocardiographic recordings were acquired for each subject. Oxygen saturation via pulse oximetry was recorded continuously. Blood pressure was measured with a manual sphygmomanometer every 3 to 4 minutes. Capillary lactate was measured prior to exercise and again at peak effort. Following 2 minutes of unloaded cycling, the work rate was increased to 20W. This was followed by a 10W/min ramp. Predetermined conditions for termination of the test included arrhythmia, chest pain, hypotensive response during exercise or an excessive hypertensive response during the test (>250/115 mm Hg). Otherwise, participants were encouraged to reach their maximal work rate. Following peak effort, participants performed a further 3 minutes of unloaded cycling.
Maximal voluntary ventilation (MVV) was calculated by multiplying $FEV_1$ by 40. Maximum oxygen uptake ($VO_2$ max) was predicted using the Wasserman weight algorithm.[40] Maximum heart rate was predicted using the equation 220 – age in years.[41] The V-slope method was used to identify the anaerobic threshold (AT). This was normalized to peak $VO_2$ as ($VO_2$ at AT/predicted maximal $VO_2$) x 100. All data points were averaged every 20 seconds.

**Six minute walk test (6MWT)**
The walk test was performed in an indoor premarked corridor. Borg scale, heart rate and oxygen saturation were measured immediately before and after the test. Participants were instructed to cover the maximal distance within 6 minutes by walking from one end of the corridor to the other, and to avoid talking during the test.[42]

**Questionnaires**

**Patient health questionnaire (PHQ-9)**
The Patient Health Questionnaire depression module (PHQ-9),[43] a multipurpose instrument for screening, diagnosing and monitoring depression, was administered to assess for evidence of depression.[44] Scores of 5, 10, and 15 were used as cut-offs mild, moderate, and severe depression, respectively.

**General Anxiety Disorder Questionnaire (GAD-7)**
The Generalised Anxiety Disorder Assessment (GAD-7),[44] a seven-item instrument, was used to screen for and measure the severity of generalised anxiety disorder among participants. Scores of 5, 10, and 15 were used as cut-offs for mild, moderate, and severe anxiety, respectively.

**Short form (SF-36)**
The SF-36 survey assesses health status and quality of life. Scores for each domain (a total of 8) were calculated using the survey scoring instructions; all scores range from 0 (worst) to 100 (best), with higher scores indicating better quality of life.[45]

**Montreal Cognitive Assessment (MoCA) Tool**
Cognitive function was assessed by using the MoCA,[46] with scores ranging from 0 to 30 points. A score of 18-25 is indicative of mild cognitive impairment, 10-17 is moderate, and <10 is severe.[46]

**Medical Research Council scale (MRC)**
The MRC dyspnoea scale[47] was used assess the effects of breathlessness on daily activities. This scale was used to measure perceived respiratory disability on a scale of 1 (no breathlessness) to 5 (complete incapacity).

**Dyspnoea-12**
The dyspnoea-12 questionnaire was used to assess both the physical and affective aspects of breathlessness. This consisted of 12 questions, each assessed on a scale of 0 (none) to 3 (severe). The total score consisted of the sum of each individual score, and ranged from 0 to 36, with a higher score reflecting more severe symptoms.[48]

**Fatigues Severity Scale**
The Fatigue Severity Scale (FSS) is a symptom-specific outcome scale aimed at providing a standardised means of describing health state.[49] The FSS comprises of nine statements, describing the severity and impact of fatigue, with a scale of possible responses ranging from 1 (“strongly disagree”) to 7 (“strongly agree”). FSS was reported as the mean score over the nine items; a higher mean score (≥4) indicates greater severity.

**Table 1. Blood test results for patient with COVID-19 during admission, at follow-up and control participants.**

| | COVID (admission) | COVID (post-discharge) | CONTROL | p-value |
|---|---|---|---|---|
| **Haematology and Coagulation** | | | | |
| White cell count, x$10^9$ / L | 6·5 (5·0 - 8·1) | 6·5 (1·8) | 6·7 (1·6) | 0·72 |
| <4 | 6/58 (10·3%) | 5/57 (8·8%) | 0/30 (0%) | |
| 4 - 10 | 45/58 (77·6%) | 52/57 (91·2%) | 28/30 (93·3%) | |
| >10 | 7/58 (12·1%) | 0/57 (0%) | 2/30 (6·7%) | |
| Neutrophil count, x$10^9$ / L | 5·2 (3·5 - 6·6) | 3·6 (2·9 - 4·6) | 3·9 (2·8 - 4·3) | 0·65[+] |
| Lymphocyte count, x$10^9$ / L | 0·9 (0·7 - 1·3) | 1·8 (1·6 - 2·3) | 1·9 (1·6 - 2·5) | 0·91[+] |
| <1·0 | 27/58 (46·6%) | 0/57 (0%) | 0/30 (0%) | |
| Haemoglobin, g/L | 141·0 (125·5 - 150·5) | 135·4 (13·2) | 139·0 (14·4) | 0·25 |
| Platelet count, x$10^9$ / L | 207·5 (168·8 - 259·5) | 261·0 (213·5 - 285·5) | 269·0 (220·0 - 292·0) | 0·63[+] |
| <100 | 1/58 (1·7%) | 0/57 (0%) | 0/30 (0%) | |
| PT, s | 10·5 (10·3 - 10·9) | 10·4 (10·2 - 10·8) | 10·5 (10·3 - 10·8) | 0·50[+] |
| APTT, s | 24·1 (2·5) | 25·3 (23·5 - 26·6) | 24·4 (22·5 - 25·8) | 0·084[+] |
| D-dimer, mg/L | 780·0 (636·0 - 1490·0) | 418·0 (253·8 - 829·3) | 337·0 (227 - 498·75) | 0·054[+] |
| **Hepatic panel** | | | | |
| Total bilirubin, mmol/L | 10·0 (7·0 - 13·8) | 10·0 (6·8 - 14·0) | 8·0 (7·0 - 11·5) | 0·51[+] |
| ALT, IU/L | 34·0 (22·3 - 62·8) | 243·5 (18·8 - 39·0) | 23·5 (16·0 - 28·0) | 0·093[+] |
| >135 IU/L (>3xULN) | 24/56 (42·9%) | 1/58 (1·7%) | 0/30 | 1·00[ε] |
| Alk Phos , IU/L | ·· | 72·0 (60·0 - 85·5) | 65·5 (55·8 - 80·3) | 0·21[+] |
| >260 IU/L (>2xULN) | ·· | 0/58 | 0/30 | |
| AST, IU/L | ·· | 23·0 (18·0 - 28·0) | 21·0 (18·0 - 27·0) | 0·36[+] |
| >126 IU/L (>3xULN) | ·· | 0/55 | 0/25 | |
| GGT , IU/L | ·· | 33·0 (21·8 - 52·3) | 29·0 (18·5 - 47·5) | 0·25[+] |
| >80 IU/L (>2xULN) | .. | 6/54 (11·1%) | 1/25 (4%) | 0·42[ε] |
| **Renal function and electrolytes** | | | | |
| Potassium, mmol/L | 3·8 (3·7 - 4·1) | 3·9 (0·3) | 3·9 (0·3) | 0·92 |

| | | | | |
|---|---|---|---|---|
| Sodium, mmol/L | 136·0 (2·9) | 141·0 (139·0 - 141·3) | 140·0 (139·0 - 141·0) | 0·12+ |
| Creatinine, umol/L | 75·5 (69·0 - 91·0) | 69·5 (60·0 - 79·3) | 79·0 (63·0 - 89·0) | 0·16+ |
| ≤133 | 55/58 (94·8%) | 57/58 (98·3%) | 30/30 (100%) | |
| >133 | 3/58 (5·2%) | 1/58 (1·7%) | 0/30 (0%) | |
| eGFR | | | | |
| ≥60 | 52/58 (89·7%) | 55/58 (94·8%) | 30/30 (100%) | 0·70ε |
| 45-59 | 3/58 (5·2%) | 1/58 (1·7%) | 0 (0%) | |
| 30-44 | 2/58 (3·4%) | 2/58 (3·4%) | 0 (0%) | |
| 15-29 | 1/58 (1·7%) | 0/58 (0%) | 0 (0%) | |
| <15 | 0/58 (0%) | 0/58 (0%) | 0 (0%) | |
| **Inflammatory markers** | | | | |
| C-reactive protein, mg/L | 119·1 (75·9 - 185·5) | 2·0 (0·9 - 5·0) | 1·2 (0·70 - 2·60) | 0·058+ |
| Procalcitonin, ug/L | ·· | 0·020 (0·020 - 0·040) | 0·02 (0·02 - 0·03) | 0·80+ |
| **Heart failure and cardiac injury** | | | | |
| NT-proBNP, ng/L | ·· | 56·8 (32·3 - 113·6) | 48·1 (23·0 - 88·4) | 0·22+ |
| Troponin I, ng/L | ·· | 2·0 (2·0 - 3·0) | 2·0 (2·0 - 3·0) | 0·49+ |
| **Peak values during admission** | | | | |
| ALT, IU/L | 70·0 (33·0 - 167·0) | | | |
| >135 IU/L (>3xULN) | 18/57 (31·2%) | | | |
| Alk Phos, IU/L | 91·0 (70·0 - 150·0) | | | |
| >260 IU/L (>2xULN) | 4/57 (7·0%) | | | |
| AST, IU/L | 49·0 (35·5 - 94·8) | | | |
| >126 IU/L (>3xULN) | 4/24 (16·7%) | | | |
| Total bilirubin, mmol/L | 11·0 (8·0 - 16·0) | | | |
| Creatinine, umol/L | 77·0 (69·0 - 93·0) | | | |
| C-reactive protein, mg/L | 185·8 (118·5 - 252·4) | | | |
| D-dimer, mg/L | 1607·5 (634·0 - 4381·8) | | | |
| Ferritin, ug/L | 594·5 (326·8 - 1501·9) | | | |
| Troponin I, ng/L | 6·5 (3·3 - 15·5) | | | |

| | |
|---|---|
| White cell count, $x10^9$ / L | 8·5 (6·7 - 13·1) |
| **Trough values during admission** | |
| eGFR, ml/min/1·73m$^2$ | 87·0 (69·8 - 90·0) |
| ≥60 | 46/58 (79·3%) |
| 45-59 | 7/58 (12·1%) |
| 30-44 | 2/58 (3·4%) |
| 15-29 | 1/58 (1·7%) |
| <15 | 2/58 (3·4%) |
| Platelet count, $x10^9$ / L | 189·0 (160·0 - 232·0) |
| White cell count, $x10^9$ / L | 5·5 (4·6 - 7·0) |
| Lymphocyte count, $x10^9$ / L | 0·9 (0·7 - 1·3) |

Data are median (IQR) for non-parametric data and mean (SD) for parametric data, and n/N (%), where N is the total number of participants with available data. P values comparing COVID (post-discharge) and CONTROL are from independent t-test, Mann-Whitney U test ($^+$) or Fisher's exact test ($^\varepsilon$). PT = Prothrombin time. APTT = Activated partial thromboplastin time. ALT = Alanine aminotransferase. Alk Phos = Alkaline phosphatase. AST = Aspartate aminotransferase. GGT = Gamma-glutamyl transferase. eGFR = Estimated Glomerular Filtration Rate. CRP = C-reactive protein. BNP = B Type natriuretic peptide. Control subjects were matched for co-morbidities as closely as possible.

**Table 2. All MRI parameters in COVID-19 survivors and controls**

| | COVID | CONTROL | p-value |
|---|---|---|---|
| **Lung MRI** | | | |
| Lung parenchymal abnormalities, % | 32 (60·4%) | 3 (10·7%) | <0·0001$^{\varepsilon}$ |
| 0 | 21/53 (39.6%) | 25/28 (89·3%) | 0·0003$^{\varepsilon}$ |
| < 25% | 3/53 (5·7%) | 0/28 (0·0%) | |
| 25 - 50% | 8/53 (15·1%) | 2/28(7·1%) | |
| 50 - 75% | 9/53 (17·0%) | 0/28 (0·0%) | |
| > 75% | 12/53 (22·6%) | 1/28 (3·6%) | |
| **Cardiac MRI** | | | |
| **Left ventricular cine analysis** | | | |
| Left ventricular end diastolic volume, mls | 143·8 (127·3 - 165·9) | 153·3 (124·5 - 178·5) | 0·59$^{+}$ |
| Left ventricular end systolic volume, mls | 53·1 (41·5 - 71·7) | 53·1 (47·7 - 70·3) | 0·81$^{+}$ |
| Left ventricular mass (diastole), g | 116·1 (100·1 - 135·1) | 107·3 (84·3 - 138·3) | 0·39$^{+}$ |
| Left ventricular mass (indexed), g/m$^2$ | 58·9 (49·8 - 66·2) | 53·8 (48·6 - 63·6) | 0·21$^{+}$ |
| Left ventricular end diastolic volume (indexed), mls/m$^2$ | 73·3 (64·5 - 83·5) | 75·6 (63·4 - 87·5) | 0·51$^{+}$ |
| Left ventricular end systolic volume (indexed), mls/m$^2$ | 26·5 (19·5 - 35·0) | 28·9 (22·5 - 34·0) | 0·81$^{+}$ |
| Left ventricular stroke volume, mls | 89·6 (79·5 - 104·7) | 95·0 (78·4 - 116·5) | 0·59$^{+}$ |
| Left ventricular stroke volume (indexed), mls/m$^2$ | 46·7 (39·9 - 52·0) | 45·1 (40·9 - 55·6) | 0·74$^{+}$ |
| Left ventricular ejection fraction, % | 63·0 (7·72) | 63·6 (6·32) | 0·70 |
| **Right ventricular cine analysis** | | | |
| Right ventricular end diastolic volume, mls | 164·4 (36·6) | 169·3 (46·5) | 0·61 |
| Right ventricular end systolic volume, mls | 70·4 (23·6) | 72·7 (24·2) | 0·69 |
| Right ventricular mass, g | 28·8 (25·8 - 35·5) | 33·2 (23·7 - 41·8) | 0·26$^{+}$ |
| Right ventricular mass (indexed), g/m$^2$ | 14·4 (12·6 - 17·2) | 16·7 (13·9 - 19·3) | 0·19$^{+}$ |
| Right ventricular end diastolic volume (indexed), mls/m$^2$ | 81·8 (14·0) | 84·3 (18·5) | 0·51 |
| Right ventricular end systolic volume (indexed), mls/m$^2$ | 34·8 (10·0) | 36·2 (10·4) | 0·58 |
| Right ventricular stroke volume, mls | 94·0 (19·3) | 96·6 (25·6) | 0·61 |
| Right ventricular stroke volume (indexed), mls/m$^2$ | 47·0 (8·1) | 48·1 (10·0) | 0·59 |
| Right ventricular EF, % | 57·9 (7·8) | 57·6 (6·0) | 0·85 |
| **$T_1$ and $T_2$ map analysis** | | | |

| | | | |
|---|---|---|---|
| Native $T_1$ (basal myocardium), ms | 1179·7 (34·4) | 1149·3 (24) | 0·0001 |
| > 1197 ms (>2SD from control mean) | 13/50 (26·0%) | 1/28 (3·7%) | 0·015[ε] |
| Native $T_1$ (mid myocardium), ms | 1173·1 (33·6) | 1150·2 (32·4) | 0·004 |
| > 1215 ms (>2SD from control mean) | 4/51 (7·8%) | 0/28 (0%) | 0·29[ε] |
| Native $T_1$ (apical myocardium), ms | 1177·4 (44·7) | 1168·3 (53·2) | 0·42 |
| Native $T_1$ (apical myocardium) > 1275 ms (>2SD from control mean) | 1/50 (2·0%) | 1/28 (3·6%) | 1·00[ε] |
| Extracellular volume (basal myocardium), % | 30·4 (28·3 - 31·3) | 28·3 (26·8 - 31·5) | 0·12 |
| Extracellular volume (mid myocardium), % | 30·1 (27·2 - 31·4) | 29·4 (27·1 - 30·7) | 0·41[+] |
| Extracellular volume (apical myocardium), % | 28·7 (27·0 - 31·6) | 29·7 (27·2 - 31·5) | 0·51[+] |
| $T_2$ (basal myocardium), ms | 41·7 (2·2) | 41·6 (2·2) | 0·80 |
| > 46 ms (>2SD from control mean) | 3/50 (6·0%) | 1/28 (3·6%) | 1·00[ε] |
| $T_2$ (mid myocardium), ms | 41·8 (2·2) | 41·1 (2·3) | 0·21 |
| > 46 ms (>2SD from control mean) | 1/50 (2·0%) | 1/28 (3·6%) | 1·00[ε] |
| $T_2$ (apical myocardium), ms | 43·5 (3·0) | 43·7 (3·5) | 0·81[+] |
| > 51ms (>2SD from control mean) | 1/50 (2·0%) | 1/28 (3·6%) | 1·00[ε] |
| **Late gadolium enhancement analysis** | | | |
| % LGE volume enhancement, % of LV | 0·8 (0·5 - 1·9) | 0·6 (0·3 - 1) | 0·023[+] |
| Myocarditis pattern | 6/52 (11·5%) | 2/28 (7·4%) | 0·47[ε] |
| Myocardial infarction | 1/52 (1·9%) | 0(0%) | |
| LV/RV insertion point | 7/52 (13·5%) | 1/28 (3·7%) | |
| Mixed | 0 (0%) | 0 (0%) | |
| Other | 0 (0%) | 0 (0%) | |
| Pericardial effusion >10mm | 1/52 (1·9%) | 0/52 (0%) | 1·00 |
| **Liver analysis** | | | |
| $T_1$, ms | 812·3 (761·3 - 896·2) | 770·0 (708·3 - 841·3) | 0·059[+] |
| $T_2$*, ms | 17·7 (4·4) | 17·2 (3·49) | 0·60 |
| Iron-corrected liver T1[¶], ms | 861·0 (99·2) | 803·9 (106·6) | 0·019 |
| > 1016ms (>2SD from control mean) | 5/52 (9·6%) | 1/28 (3·6%) | 0·66[ε] |
| Average proton density fat fraction, % | 4·9 (3·1 - 9·5) | 3·7 (2·1 - 6·5) | 0·18[+] |

| | | | |
|---|---|---|---|
| Extracellular volume, % | 27·3 (23·0 - 31·2) | 27·3 (17·6 - 33·6) | 0·60$^{+}$ |
| **Spleen analysis** | | | |
| Volume, mls | 216·4 (179·8 - 269·4) | 204·9 (149·6 - 248·6) | 0·47$^{+}$ |
| Volume (indexed), mls | 102·9 (87·4 - 141·0) | 105·2 (78·4 - 133·7) | 0·66$^{+}$ |
| $T_1$, ms | 1332·0 (1269·0 - 1376·3) | 1304·0 (1176·5 - 1364·0) | 0·37$^{+}$ |
| $T_2$*, ms | 20·0 (15·1 - 30·7) | 23·6 (11·7 - 32·7) | 0·93$^{+}$ |
| **Renal analysis** | | | |
| **$T_1$ map anaylsis** | | | |
| Left cortex, ms | 1605·5 (93·6) | 1534·0 (74·8) | 0·001 |
| Right cortex, ms | 1591·4 (91·8) | 1514·5 (63·0) | 0.0002 |
| Average cortex, ms | 1597·5 (91·2) | 1523·1 (65·5) | 0.0003 |
| > 1652ms (>2SD from control mean)) | 15/51 (29·4%) | 0/28 (0%) | 0·001$^{\varepsilon}$ |
| Left medulla, ms | 2001·5 (82·9) | 1985·8 (76·6) | 0·41 |
| Right medulla, ms | 1983·6 (72·6) | 1990·7 (74·4) | 0·68 |
| Average medulla, ms | 1992·6 (71·6) | 1988·8 (73·7) | 0·82 |
| Left corticomedullary differentiation, ms | 396·5 (97·6) | 451·8 (75·0) | 0·011 |
| Right corticomedullary differentiation, ms | 392·4 (349 - 444·9) | 478·7 (423·6 - 510·5) | <0·0001$^{+}$ |
| Average corticomedullary differentiation, ms | 385·4 (335·8 - 456·4) | 470·8 (431·5 - 496·3) | 0·0002$^{+}$ |
| **Blood oxygen level dependent imaging** | | | |
| Left kidney radial profile gradient, Hz/% | 0·18115 (0·07510) | 0·17740 (0·06725) | 0·85 |
| Right kidney radial profile gradient, Hz/% | 0·16684 (0·09202) | 0·17438 (0·07041) | 0·89 |
| Average kidney radial profile gradient, Hz/% | 0·16756 (0·06938) | 0·17165 (0·06559) | 0·51 |
| **Brain analysis** | | | |
| **$T_1$ volumes (Sienax)** | | | |
| Head size scaling | 1·3 (1·3 - 1·4) | 1·3 (1·2 - 1·4) | 0·42$^{\gamma}$ |
| Peripheral gray matter volume (normalised), $mm^3$ | 627811·3 (603839·0-664594·8) | 650373·3 (616362·8-670797·4) | 0·25$^{\gamma}$ |
| Peripheral gray matter volume, $mm^3$ | 476350 (442739·6-507886·6) | 492021·8 (460985·1-528880·0) | 0·15$^{\gamma}$ |
| Cerebrospinal fluid (normalised), $mm^3$ | 35334 (26810·8-50216·4) | 34998·7 (27688·2-54755·5) | 0·43$^{\gamma}$ |

| | | | |
|---|---|---|---|
| Cerebrospinal fluid, mm$^3$ | 25502·2 (19964·1-38665·7) | 28163·5 (20168·6-45052·7) | 0·37[γ] |
| Gray matter volume (normalised), mm$^3$ | 785448·6 (754636·9-825737·9) | 801955·7 (763848·3-831534·2) | 0·43[γ] |
| Gray matter volume, mm$^3$ | 589913·2 (557440·5-624603·5) | 609307·8 (574621·1-660878·3) | 0·49[γ] |
| White matter volume (normalised), mm$^3$ | 749563·9 (718930·5-785434·2) | 752374 (737043·6-771490·9) | 0·75[γ] |
| White matter volume, mm$^3$ | 573062·1 (537902·4-610030·0) | 576954·4 (520135·8-625013·0) | 0·98[γ] |
| Brain (normalised), mm$^3$ | 1558166 (1480596-1596700) | 1564271 (1508934-1610240) | 0·87[γ] |
| Brain, mm$^3$ | 1163509 (1100464-1226565) | 1186262 (1089109-1285950) | 0·56[γ] |
| **$T_1$ volumes (First)** | | | |
| Left thalamus, mm$^3$ | 7784 (7201·0-8202·8) | 7837 (7265·8-8564·5) | 0·61[γ] |
| Right thalamus, mm$^3$ | 7661 (6963·2-8039·0) | 7704 (7082·0-8246·0) | 0·80[γ] |
| Left caudate, mm$^3$ | 3335 (3080·2-3692·2) | 3443 (3301·8-3556·5) | 0·70[γ] |
| Right caudate, mm$^3$ | 3464 (3204·8-3808·0) | 3597 (3338·8-3768·2) | 0·80[γ] |
| Left putamen, mm$^3$ | 4814 (4530·0-5106·2) | 5029 (4582·5-5443·2) | 0·86[γ] |
| Right putamen, mm$^3$ | 4825 (4488·2-5175·5) | 4933 (4528·8-5542·8) | 0·79[γ] |
| Left pallidum, mm$^3$ | 1776 (1630·8-1884·8) | 1717 (1536·8-1890·8) | 0·74[γ] |
| Right pallidum, mm$^3$ | 1836 (1698·5-1947·8) | 1882 (1616·0-2045·5) | 0·72[γ] |
| Left hippocampus, mm$^3$ | 3727 (3474·0-3906·2) | 3846 (3691·5-4120·8) | 0·17[γ] |
| Right hippocampus, mm$^3$ | 3746 (3487·0-4053·5) | 4039 (3709·2-4230·8) | 0·067[γ] |
| Left amygdala, mm$^3$ | 1346 (1120·5-1506·5) | 1334 (1228·5-1491·5) | 0·69[γ] |
| Right amygdala, mm$^3$ | 1331 (1176·8-1512·0) | 1317 (1209·8-1493·8) | 0·88[γ] |
| Left accumbens, mm$^3$ | 542 (477·2-583·8) | 554 (515·5-622·2) | 0·75[γ] |
| Right accumbens, mm$^3$ | 423 (338·2-481·0) | 461 (356·2-538·0) | 0·21[γ] |
| Brain stem and 4th ventricle, mm$^3$ | 22082·0 (20259·5 - 23365·0) | 22595·0 (19518·5 - 24308·5) | 0·58[γ] |
| **$T_2$-FLAIR volumes** | | | |
| White matter hyperintensities, mm$^3$ | 2305·0 (1402·0-4021·0) | 1457·0 (654·2-2700·5) | 0·085[γ] |
| Periventricular white matter hyperintensities, mm$^3$ | 1884·0 (1172·0-3303·0) | 1305·0 (525·0-2284·8) | 0·066[γ] |
| Deep white matter hyperintensities, mm$^3$ | 330·5 (141·0-863·0) | 213·0 (83·5-416·8) | 0·20[γ] |
| **Susceptibility weighted imaging, $T_2$*** | | | |

| | | | |
|---|---|---|---|
| Left thalamus, ms | 44·2 (42·0-46·1) | 42·8 (39·9-45·3) | 0·047γ |
| Right thalamus, ms | 43·9 (41·7-45·8) | 42·4 (40·2-45·0) | 0·034γ |
| Left caudate, ms | 41·4 (38·5-43·5) | 42·9 (39·6-44·1) | 0·71γ |
| Right caudate, ms | 41·7 (37·8-44·4) | 42·1 (39·7-43·1) | 0·37γ |
| Left putamen, ms | 38 (32·9-40·2) | 36·9 (33·2-39·1) | 0·32γ |
| Right putamen, ms | 36·7 (33·1-40·1) | 35·6 (33·6-38·1) | 0·24γ |
| Left pallidum, ms | 27·6 (24·4-30·0) | 26·7 (25·9-28·6) | 0·54γ |
| Right pallidum, ms | 27.0 (24·0-29·6) | 26·4 (25·3-28·7) | 0·64γ |
| Left hippocampus, ms | 43·5 (40·9-45·9) | 45·4 (43·9-47·7) | 0·038γ |
| Right hippocampus, ms | 44·1 (41·9-46·0) | 45 (42·8-48·7) | 0·21γ |
| Left amygdala, ms | 44·2 (41·3-47·6) | 46 (41·6-47·2) | 0·60γ |
| Right amygdala, ms | 44·3 (42·1-46·6) | 46·6 (43·9-48·3) | 0·75γ |
| Left accumbens, ms | 40·0 (36·5-44·5) | 39·6 (35·8-43·6) | 0·42γ |
| Right accumbens, ms | 41·0 (38·1-44·2) | 40·5 (37·1-43·2) | 0·38γ |
| **Susceptibility weighted imaging, quantitative susceptibility mapping** | | | |
| Left thalamus, ppb | -3·2 (-7·5--0·4) | -3·5 (-5·6--1·1) | 0·76γ |
| Right thalamus, ppb | -1·4 (-3·7-2·1) | -1·4 (-4·4-0·5) | 0·54γ |
| Left caudate, ppb | 27·1 (23·9-38·3) | 28·7 (24·7-32·8) | 0·99γ |
| Right caudate, ppb | 26·8 (22·2-36·5) | 28·3 (25·2-34·3) | 0·12γ |
| Left putamen, ppb | 16·3 (11·0-23·9) | 18·6 (13·4-23·6) | 0·26γ |
| Right putamen, ppb | 20·4 (13·5-26·7) | 18·1 (11·2-25·0) | 0·96γ |
| Left pallidum, ppb | 73·6 (59·5-86·6) | 80 (68·1-87·9) | 0·72γ |
| Right pallidum, ppb | 72·5 (58·2-85·5) | 76·2 (62·4-85·1) | 0·58γ |
| Left hippocampus, ppb | 5·6 (3·0-7·6) | 5·9 (3·6-9·4) | 0·57γ |
| Right hippocampus, ppb | 6·0 (3·5-9·7) | 5·7 (2·6-7·0) | 0·39γ |
| Left amygdala, ppb | 2·6 (-4·7-6·8) | 5·9 (0·3-9·0) | 0·41γ |
| Right amygdala, ppb | 2·7 (-2·1-6·5) | 6.0 (-1·7-11·8) | 0·40γ |
| Left accumbens, ppb | -0·1 (-5·4-1·8) | 0·0 (-4·7-2·2) | 1·00γ |

| | | | |
|---|---|---|---|
| Right accumbens, ppb | 0·0 (-6·0-0·6) | 0·0 (-9·1-3·3) | 0·62[γ] |
| **Arterial spin labelling, mean measures in gray matter** | | | |
| Arterial arrival time, s | 1·4 (1·3-1·5) | 1·4 (1·3-1·4) | 0·68[γ] |
| Cerebral blood flow (mean perfusion), ml/100g/min | 54·4 (48·4-58·8) | 53·1 (49·8-61·0) | 0·80[γ] |
| **Diffusion weighted imaging** | | | |
| Middle cerebellar peduncle, x10$^{-6}$ mm$^2$/s | 695·0 (679·0-708·5) | 686·0 (677·0-700·8) | 0·57[γ] |
| Pontine crossing tract, x10$^{-6}$ mm$^2$/s | 697·0 (680·0-727·0) | 701·0 (663·0-720·8) | 0·75[γ] |
| Genu of corpus callosum , x10$^{-6}$ mm$^2$/s | 783·0 (762·5-808·0) | 784·0 (764·2-806·2) | 0·87[γ] |
| Body of corpus callosum, x10$^{-6}$ mm$^2$/s | 770·0 (744·0-798·5) | 760·0 (746·0-779·2) | 1·00[γ] |
| Splenium of corpus callosum, x10$^{-6}$ mm$^2$/s | 769·0 (747·0-779·8) | 753·0 (739·8-779·2) | 0·28[γ] |
| Fornix, x10$^{-6}$ mm$^2$/s | 1288·0 (1081·5-1574·2) | 1298·0 (1094·0-1609·8) | 0·51[γ] |
| Right corticospinal Tract, x10$^{-6}$ mm$^2$/s | 690·0 (658·8-718·0) | 678·0 (648·2-706·2) | 0·28[γ] |
| Left corticospinal Tract, x10$^{-6}$ mm$^2$/s | 693·0 (669·5-724·5) | 673·0 (637·0-701·8) | 0·088[γ] |
| Right medial lemniscus, x10$^{-6}$ mm$^2$/s | 720·0 (694·2-755·8) | 714·0 (681·0-754·5) | 0·91[γ] |
| Left medial lemniscus, x10$^{-6}$ mm$^2$/s | 707·0 (681·5-724·5) | 695·0 (665·2-728·8) | 0·67[γ] |
| Inferior cerebellar peduncle, x10$^{-6}$ mm$^2$/s | 767·0 (737·8-785·0) | 753·0 (731·0-769·5) | 0·28[γ] |
| Superior cerebellar peduncle, x10$^{-6}$ mm$^2$/s | 729·0 (712·2-744·2) | 719·0 (710·0-735·2) | 0·55[γ] |
| Right cerebellar peduncle, x10$^{-6}$ mm$^2$/s | 786·0 (771·0-818·2) | 794·0 (761·8-822·0) | 0·47[γ] |
| Left cerebellar peduncle, x10$^{-6}$ mm$^2$/s | 763·0 (737·5-780·8) | 756·0 (744·5-780·2) | 0·66[γ] |
| Right cerebral peduncle, x10$^{-6}$ mm$^2$/s | 736·0 (720·2-756·5) | 737·0 (718·8-761·0) | 0·82[γ] |
| Left cerebral peduncle, x10$^{-6}$ mm$^2$/s | 703·0 (682·0-726·0) | 703·0 (690·2-724·2) | 0·64[γ] |
| Anterior limb of right internal capsule, x10$^{-6}$ mm$^2$/s | 742·0 (725·0-767·0) | 737·0 (726·5-750·8) | 0·38[γ] |
| Anterior limb of the left internal capsule, x10$^{-6}$ mm$^2$/s | 719·0 (696·2-752·8) | 727·0 (698·5-742·2) | 0·64[γ] |
| Posterior limb of right internal capsule, x10$^{-6}$ mm$^2$/s | 711·0 (703·0-735·5) | 713·0 (701·8-723·5) | 0·69[γ] |
| Posterior limb of the left internal capsule, x10$^{-6}$ mm$^2$/s | 683·0 (668·0-697·2) | 676·0 (658·5-693·5) | 0·52[γ] |
| Retrolenticular part of right internal capsule, x10$^{-6}$ mm$^2$/s | 825·0 (806·0-852·0) | 820·0 (804·0-839·2) | 0·52[γ] |
| Retrolenticular part of left internal capsule, x10$^{-6}$ mm$^2$/s | 780·0 (762·0-803·5) | 778·0 (742·0-793·0) | 0·18[γ] |
| Right anterior corona radiata, x10$^{-6}$ mm$^2$/s | 762·0 (743·2-785·2) | 751·0 (738·5-761·8) | 0·27[γ] |

| | | | |
|---|---|---|---|
| Left anterior corona radiata, x10$^{-6}$ mm$^2$/s | 760·0 (741·2-792·5) | 763·0 (747·2-773·2) | 0·91[ɣ] |
| Right superior corona radiata, x10$^{-6}$ mm$^2$/s | 721·0 (695·5-738·8) | 702·0 (692·8-715·8) | 0·23[ɣ] |
| Left superior corona radiata, x10$^{-6}$ mm$^2$/s | 711·0 (681·0-729·5) | 689·0 (676·0-701·0) | 0·33[ɣ] |
| Right posterior corona radiata, x10$^{-6}$ mm$^2$/s | 834·0 (812·5-871·8) | 825·0 (800·5-854·2) | 0·92[ɣ] |
| Left posterior corona radiata, x10$^{-6}$ mm$^2$/s | 799·0 (777·0-824·5) | 784·0 (769·0-802·2) | 0·13[ɣ] |
| Right posterior thalamic radiation, x10$^{-6}$ mm$^2$/s | 842·0 (804·5-871·2) | 813·0 (787·0-832·8) | 0·19[ɣ] |
| Left posterior thalamic radiation, x10$^{-6}$ mm$^2$/s | 831·0 (814·5-851·5) | 811·0 (792·2-828·8) | 0·042[ɣ] |
| Right sagittal stratum, x10$^{-6}$ mm$^2$/s | 840·0 (799·5-863·5) | 813·0 (789·2-828·5) | 0·022[ɣ] |
| Left sagittal stratum, x10$^{-6}$ mm$^2$/s | 789·0 (776·5-814·0) | 787·0 (767·2-791·5) | 0·078[ɣ] |
| Left and right sagittal stratum, x10$^{-6}$ mm$^2$/s | 810·0 (791.0-834.0) | 791·5 (779·5-808·8) | 0·02[ɣ] |
| Right external capsule, x10$^{-6}$ mm$^2$/s | 783·0 (766·0-816·2) | 777·0 (755·8-796·0) | 0·224[ɣ] |
| Left external capsule, x10$^{-6}$ mm$^2$/s | 765·0 (744·0-794·0) | 752·0 (734·5-776·5) | 0·107[ɣ] |
| Right cingulum cingulate gyrus, x10$^{-6}$ mm$^2$/s | 746·0 (720·2-768·8) | 736·0 (722·5-757·0) | 0·46[ɣ] |
| Left cingulum cingulate gyrus, x10$^{-6}$ mm$^2$/s | 745·0 (721·2-774·0) | 745·0 (710·0-764·2) | 0·55[ɣ] |
| Right cingulum hippocampus, x10$^{-6}$ mm$^2$/s | 783·0 (746·2-802·8) | 773·0 (745·2-807·8) | 0·88[ɣ] |
| Left cingulum hippocampus, x10$^{-6}$ mm$^2$/s | 740·0 (711·5-784·2) | 758·0 (736·8-786·0) | 0·50[ɣ] |
| Right fornix cres and stria terminalis, x10$^{-6}$ mm$^2$/s | 884·0 (859·2-934·5) | 895·0 (859·0-919·0) | 0·88[ɣ] |
| Left fornix cres and stria terminalis, x10$^{-6}$ mm$^2$/s | 795·0 (766·8-830·8) | 788·0 (752·0-818·2) | 0·39[ɣ] |
| Right superior longitudinal fasciculus , x10$^{-6}$ mm$^2$/s | 729·0 (708·0-754·8) | 717·0 (708·0-741·2) | 0·38[ɣ] |
| Left superior longitudinal fasciculus, x10$^{-6}$ mm$^2$/s | 697·0 (679·0-711·8) | 680·0 (669·0-699·0) | 0·069[ɣ] |
| Right superior fronto-occipital fasciculus, x10$^{-6}$ mm$^2$/s | 720·0 (673·8-748·5) | 693·0 (664·5-707·2) | 0·30[ɣ] |
| Left superior fronto-occipital fasciculus, x10$^{-6}$ mm$^2$/s | 681·0 (636·5-711·5) | 666·0 (648·0-718·8) | 0·73[ɣ] |
| Right uncinate fasciculus, x10$^{-6}$ mm$^2$/s | 758·0 (719·5-797·8) | 747·0 (709·5-791·0) | 0·31[ɣ] |
| Left uncinate fasciculus, x10$^{-6}$ mm$^2$/s | 747·0 (718·0-799·0) | 731·0 (712·8-783·8) | 0·39[ɣ] |
| Right tapetum, x10$^{-6}$ mm$^2$/s | 881·0 (824·2-926·8) | 834·0 (818·5-895·5) | 0·27[ɣ] |
| Left Tapetum, x10$^{-6}$ mm$^2$/s | 849·0 (804.5-953.2) | 830·0 (784.2-902.0) | 0.43 |

Data are mean (SD), median (IQR) and n/N (%), where N is the total number of participants with available data. P values from independent t-test, Mann-Whitney U test ([+]), or Fisher's exact test ([ε]). Brain image derived phenotypes measurements (IDPs) were guassianised and deconfounded for typical brain confounders. P values for brain (ˠ) from gaussianised deconfounded model. Ppb = Parts per billion, change in magnetic susceptibility. ml/100g/min = millilitres of blood per 100g of tissue per minute. FLAIR = Fluid attenuated inversion recovery. LV =left ventricle. LGE = Late gadolinium enhancement. RV = right ventricle. Control subjects were matched for co-morbidities as closely as possible. ¶An in-house algorithm was used to calculate iron-corrected T1, so these values cannot be compared to the *LiverMultiScan* cT1[35].

**Table 3. Clinical assessment of Brain MRI scans.**

| | COVID | CONTROL | p-value |
|---|---|---|---|
| Brain abnormalities on clinical assessment | 13/54 (24·1%) | 6/28 (20·7%) | 0·73 |
| No abnormality | 41/54 (75·9%) | 23/28 (78·6%) | |
| White matter hyperintensities | 5/54 (9·3%) | 3/28 (10·7%) | |
| Small vessel disease | 5/54 (9·3%) | 3/28 (10·7%) | |
| Olfactory cortical enhancement | 1/54 (1·9%) | 0/28 (0%) | |
| Haemorrhagic or ischaemic abnormalities | 2/54 (3·7%) | 0/28 (0%) | |

Data are n/N (%), where N is the total number of participants with available data. P value from Fisher's exact test.

**Table 4. Correlation matrix between inflammatory markers at 2-3 months and severity of illness during hospital admission, imaging markers of multiorgan injury, functional capacity, depression and anxiety symptoms in COVID-19 patients and controls.**

| COVID-19 | White cell count | Neutrophil count | Monocyte | Lymphocyte | C-reactive protein | Pro-calcitonin |
|---|---|---|---|---|---|---|
| Clinical Severity (WHO ordinal scale) | 0·502 | 0·478 | 0·402 | 0·185 | 0·283 | 0·482 |
| Periventricular white matter hyperintensities | 0·466 | 0·364 | 0·262 | 0·322 | -0·004 | 0·25 |
| Average right thalamic $T_2$ | 0·287 | 0·341 | 0·100 | 0·066 | 0·130 | -0·104 |
| Average left thalamic $T_2$ | 0·113 | 0·088 | 0·159 | 0·098 | 0·028 | -0·12 |
| Myocardial basal native $T_1$ | 0·253 | 0·321 | 0·140 | -0·044 | 0·521 | 0·319 |
| Myocardial mid native $T_1$ | 0·275 | 0·271 | -0·027 | 0·136 | 0·445 | 0·233 |
| Average renal cortical native $T_1$ | 0·294 | 0·295 | 0·297 | 0·100 | 0·437 | 0·417 |
| Average renal corticomedullary differentiation | 0·048 | -0·003 | -0·103 | 0·194 | -0·299 | -0·403 |
| Iron-corrected liver $T_1$ | 0·600 | 0·614 | 0·452 | 0·249 | 0·481 | 0·484 |
| VE/$VCO_2$ slope (CPET) | 0·326 | 0·331 | 0·394 | -0·020 | 0·262 | 0·446 |
| Six minute walk distance | -0·239 | -0·241 | -0·321 | -0·083 | -0·371 | -0·322 |
| Depression (PHQ-9) score | 0·173 | 0·189 | 0·484 | -0·054 | 0·16 | 0·082 |
| Anxiety (GAD-7) score | 0·072 | 0·094 | 0·246 | -0·063 | 0·252 | 0·1 |
| **CONTROL** | **White cell count** | **Neutrophil** | **Monocyte** | **Lymphocyte** | **C-reactive protein#** | **Pro-calcitonin** |
| Periventricular white matter hyperintensities | 0·127 | 0·223 | -0·071 | -0·081 | -0·042 | 0·186 |
| Average right thalamic $T_2$ | -0·081 | -0·096 | -0·239 | 0·092 | -0·182 | -0·237 |
| Average left thalamic $T_2$ | -0·446 | -0·413 | -0·502 | -0·085 | -0·339 | 0·046 |
| Myocardial basal native $T_1$ | 0·168 | 0·135 | 0·058 | 0·158 | 0·332 | 0·149 |
| Myocardial mid native $T_1$ | 0·068 | 0·084 | -0·174 | 0·081 | 0·226 | -0·209 |
| Average renal cortical native $T_1$ | 0·234 | 0·25 | 0·221 | 0·062 | 0·24 | 0·176 |
| Average renal corticomedullary differentiation | -0·18 | -0·096 | -0·343 | -0·155 | -0·145 | -0·366 |
| Iron corrected liver $T_1$ | 0·499 | 0·44 | 0·5 | 0·285 | 0·691 | 0·229 |
| VE/$VCO_2$ slope (CPET) | -0·126 | -0·289 | -0·095 | 0·299 | 0·136 | 0·41 |
| Six minute walk distance | -0·213 | -0·076 | -0·374 | -0·325 | -0·566 | -0·121 |

| | | | | | | |
|---|---|---|---|---|---|---|
| Depression (PHQ-9) score | -0·013 | -0·054 | 0·085 | -0·028 | 0·26 | 0·355 |
| Anxiety (GAD-7) score | 0·199 | 0·128 | 0·164 | 0·033 | 0·18 | 0·293 |
| **Correlation coefficient colour scale** | -1 -0.7 | -0.3 | 0 | 0.3 | 0.7 | 1 |

COVID-19 = Coronavirus disease. WHO = world health organization. CPET = cardiopulmonary exercise test. VE/VCO2 = Ventilatory equivalent for carbon dioxide. PHQ-9 = Patient health questionnaire. GAD-7 = Generalised Anxiety Disorder Questionnaire. Pearson’s correlation coefficients are displayed for parametric data and spearman’s correlation coefficients for non-parametric correlations.

**Table 5. Correlation matrix between severity of acute illness and imaging markers of inflammation and exercise tolerance in COVID-19 patients.**

| COVID-19 | White matter hyperintensity lesions | Brain Left thalamus $T_2$* | Brain Right Thalamus $T_2$* | Cardiac base $T_1$ | Cardiac mid $T_1$ | Iron-corrected Liver $T_1$ | Renal cortical $T_1$ | Lung injury | $VE/VCO_2$ slope |
|---|---|---|---|---|---|---|---|---|---|
| WHO ordinal scale | 0.272 | -0.138 | -0.129 | 0.223 | 0.111 | 0.409 | 0.426 | 0.358 | 0.311 |
| Correlation coefficient scale | -1 | -0.7 | -0.3 | 0 | 0.3 | 0.7 | 1 | | |

COVID-19 Coronavirus disease; WHO world health organization; CPET cardiopulmonary exercise test; $VE/VCO_2$ ventilatory equivalent for carbon di-oxide. Pearson's correlation coefficients are displayed for parametric data and spearman's correlation coefficients for non-parametric data.

**Table 6. Multiorgan changes on MRI in non-critical patients versus controls**

| | Non-critical COVID (n=34) | Controls (n=28) | p-value |
|---|---|---|---|
| **Lungs** | | | |
| Parenchymal abnormalities | 15/34 (44·1%) | 3/28 (10·7%) | 0·005[ε] |
| **Myocardium** | | | |
| Native T1 (basal myocardium), ms | 1177·1 (34·9) | 1149·3 (24·0) | 0·001 |
| Native T1 (mid myocardium), ms | 1172·8 (32·6) | 1150·2 (32·4) | 0·009 |
| **Kidney** | | | |
| Average Cortex, ms | 1567·5 (79·3) | 1523·1 (65·5) | 0·023 |
| Average Corticomedullary Differentiation, ms | 423·7 (371·9 – 475·9) | 470·8 (431·5 - 496·3) | 0·040[+] |
| **Liver** | | | |
| T1, ms | 790·4 (98·5) | 778·1 (98·2) | 0·63 |
| Iron Corrected T1[¶], ms | 828·9 (85·7) | 803·9 (106·6) | 0·32 |
| **Brain[&]** | | | |
| Left thalamus susceptibility-weighted imaging T2*, ms | 44·4 (2·9) | 42·9 (3·5) | 0·032[&] |
| Right thalamus susceptibility-weighted imaging T2*, ms | 44·6 (2·6) | 42·9 (2·9) | 0·006[&] |

Data are mean (SD) and median (IQR). Non-critical defined as those patients not requiring mechanical ventilation (intubation, CPAP or BiPAP), inotropic support or renal replacement therapy. P values from independent student's t-test, Mann-Whitney U test ([+]) or Fisher's exact test ([ε]). Brain markers([&]) p-value related to gaussianised and deconfounded comparisons. [¶]An in-house algorithm was used to calculate iron-corrected T1, so these values cannot be compared to the Liver*MultiScan cT1*[35].

**Table 7. Details on missing data for variables.**

| Cohort, variable/s | Data, frequency (%) | Reason |
|---|---|---|
| **Blood values** | | |
| COVID (Admission), ALT | 56/58 (96·6%) | ALT was not measured in 2 patients during admission |
| COVID (Admission), Troponin | 38/58 (65.5%) | Troponin levels were not undertaken in 20 patients during admission |
| COVID (Post-discharge), AST | 55/58 (94·8%) | AST was not performed by the laboratory for 3 participants (lab error) |
| COVID (Post-discharge), GGT | 54/58 (93·1%) | GGT was not performed by the laboratory for 4 participants (lab error) |
| COVID (Post-discharge), Haematology data | 57/58 (98·3%) | 1 sample was insufficient for analysis, 4 specimens for full blood count assessment were clotted and recollected. |
| Lung Imaging | | |
| COVID, Lung MRI | 53/58 (91·3%) | 4 did not undergo imaging (1 due to large body habitus, 3 due to claustrophobia), 1 had limited studies due to claustrophobia during scan. |
| CONTROL, all variables | 28/30 (93·3%) | 2 did not undergo imaging due to claustrophobia. |
| **Cardiac imaging** | | |
| COVID, cardiac volumes and late gadolinium imaging | 52/58 (89·7%) | 4 did not undergo imaging (as above), 2 had limited studies without cardiac imaging due to claustrophobia during scan. |
| COVID, Native $T_1$ and $T_2$ (basal and apical myocardium) | 50/58 (86·2%) | 4 did not undergo imaging (as above), 2 had limited studies due to claustrophobia, 1 was unable to maintain a sufficient breath-hold for acquisition, 1 (base) failed quality check (QC). |
| COVID, Native $T_1$ and $T_2$ (mid myocardium) | 51/58 (87·9%) | 4 did not undergo imaging (as above), 2 had limited studies due to claustrophobia,1 was unable to maintain a sufficient breath-hold for acquisition. |
| COVID, Post-contrast $T_1$ base myocardium | 45/58 (77·6%) | 4 did not undergo imaging (as above) and 2 had limited studies due to claustrophobia, 1 was unable to maintain a sufficient breath-hold for acquisition, 3 cases failed QC in the base, haematocrit unavailable for 4 on the same day. |
| COVID, Post-contrast $T_1$ mid, apical myocardium | 46/58 (79·3%) | 4 did not undergo imaging (as above) and 2 had limited studies due to claustrophobia, 1 was unable to maintain a sufficient breath-hold for acquisition, 5 cases failed QC. |
| CONTROL, all variables | 28/30 (93·3%) | 2 did not undergo imaging due to claustrophobia. |
| CONTROL, Post-contrast $T_1$ | 26/30 (86.6%) | 2 did not undergo imaging due to claustrophobia, 2 failed QC |
| **Brain imaging** | | |
| COVID, Quantitative analysis | 51/58 (87·9%) | 4 did not undergo imaging (as above). 3 were excluded for quantitative analysis as cases failed QC at the point of image analysis. |
| **Abdominal imaging** | | |
| COVID, Kidney, Average Kidney T1 and blood oxygen level dependent imaging | 51/58 ((87·9%) | 4 did not undergo imaging (as above), 2 had limited studies without renal imaging due to claustrophobia, 1 was unable to maintain a sufficient breath-hold for acquisition. |

| | | |
|---|---|---|
| COVID, Liver, iron-corrected T1 | 52/58 (89·7%) | 4 did not undergo imaging (as above), 2 had limited studies without hepatic imaging due to claustrophobia. |
| CONTROL, all variables | 28/30 (93·3%) | 2 did not undergo imaging due to claustrophobia. |
| **Cardiopulmonary exercise testing** | | |
| COVID (Post-discharge), all variables | 51/58 (87·9%) | 3 were unable to perform the test (1 due to large body habitus, 1 due to claustrophobia on wearing mask, 1 due to acrophobia when sitting on the bicycle), 4 did not consent to the test. |
| CONTROL, all variables | 27/30 (90·0%) | 1 was unable to perform the test (due to large body habitus), 2 did not consent to the test. |
| **Pulmonary function tests** | | |
| COVID (Post-discharge), all variables | 56/58 (97·0%) | 2 did not consent to the test. |
| CONTROL, all variables | 28/30 (93·3%) | 2 did not consent to the test. |
| **Questionnaires** | | |
| COVID (Post-discharge), SF36, PHQ9, GAD7 | 57/58 (98·2%) | 1 was unable to fill in the questionnaires due to recent cerebrovascular event during admission. |
| COVID (Post-discharge), Dyspnoea-12, MRC Dyspnoea Score | 56/58 (96·5%) | 1was unable to fill in the questionnaires due to recent cerebrovascular event during admission. 1 could not complete the questionnaire due to insufficient time to complete visit. |
| COVID (Post-discharge), Fatigue Severity Score (FSS) | 55/58 (94·8%) | 1 was unable to fill in the questionnaires due to recent cerebrovascular event during admission. 2 could not complete the questionnaire due to insufficient time. |
| CONTROL, Dyspnoea-12, MRC Dyspnoea Score, Fatigue Severity Score (FSS) | 29/30 (96·6%) | 1 could not complete the questionnaire due to insufficient time to complete visit. |

Data are n/N (%), where N is the total number of participants with available data. ALT = Alanine aminotransferase. AST = Aspartate transaminase. GGT = Gamma-glutamyl transferase. GAD = Generalised anxiety disorder. PHQ-9 = Patient health questionnaire-9 assessment.

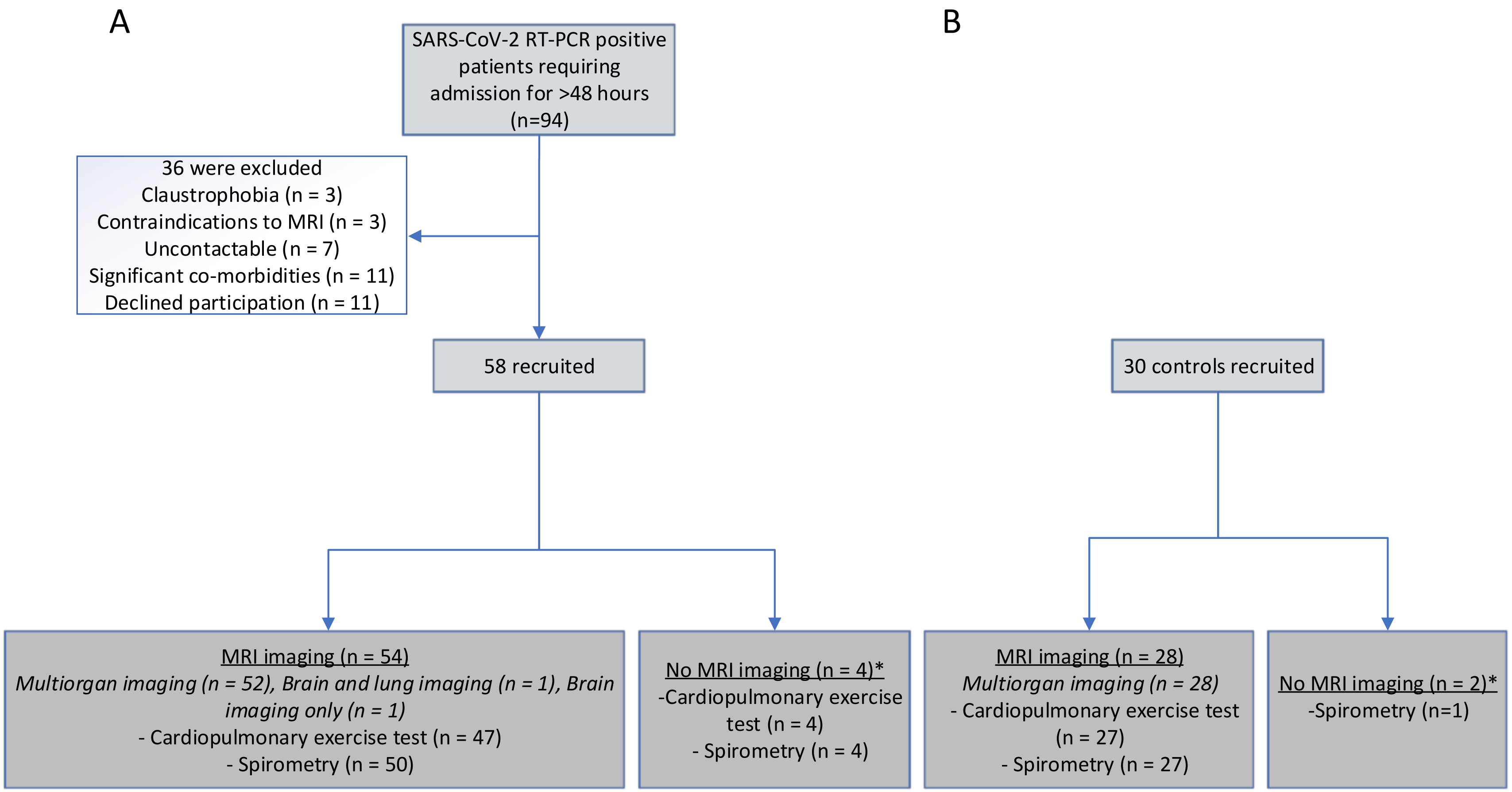

**Appendix Figure 1. Recruitment flow-chart of A) COVID-19 patients and B) Controls. *see appendix Table 7 for details**

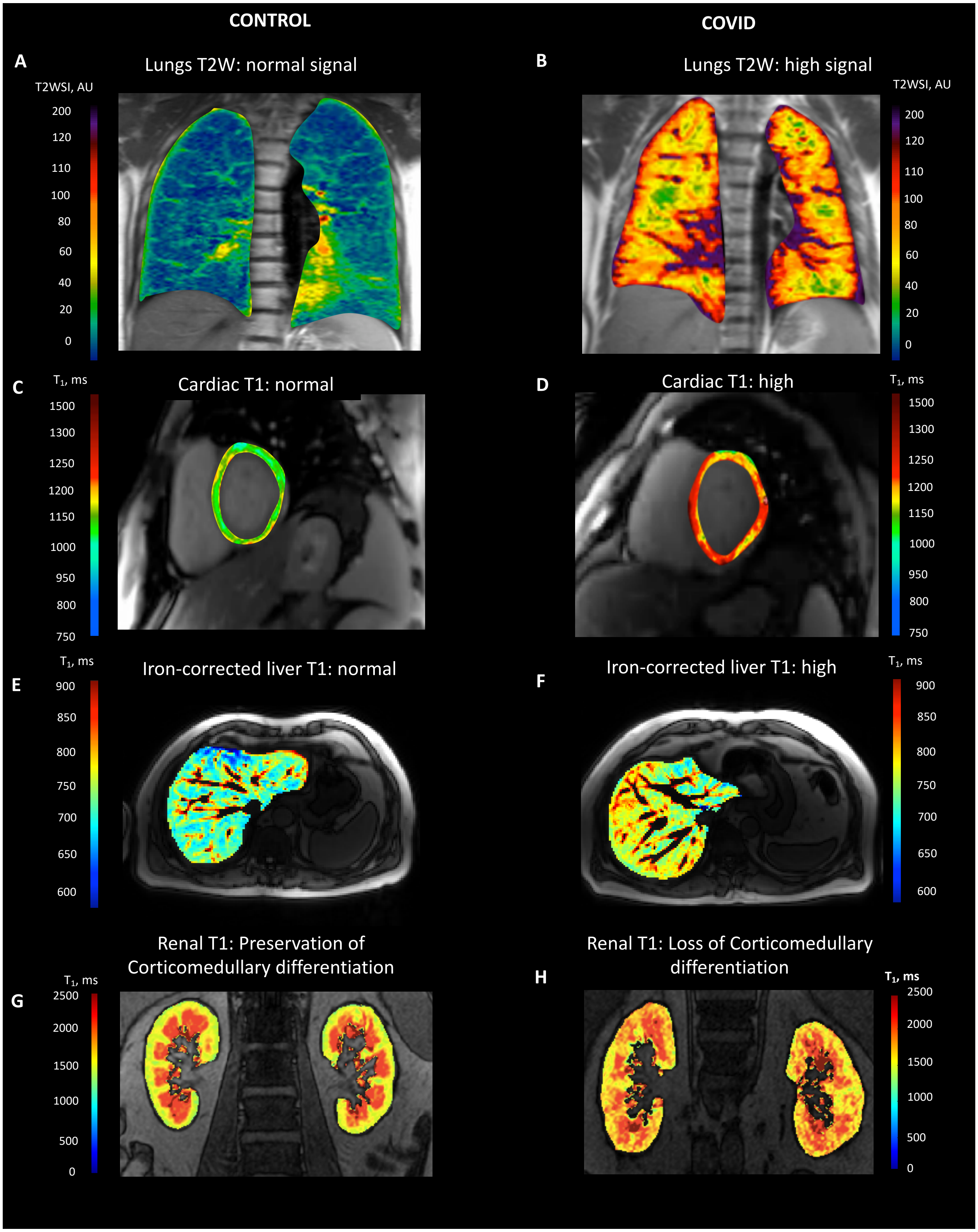

**Appendix Figure 2. Illustrative MRI images (lungs, heart, liver, kidneys) from controls (age-matched) and COVID-19 patients.**

Example cases of MRI images from uninfected controls and COVID-19 patients (matched for age) are shown here. A, B: Patients demonstrated lung parenchymal changes seen as an increase in signal (assessed qualitatively) on T2 weighted (T2w) lung MRI images compared to control subject. C, D: Myocardial abnormalities were seen as prolonged myocardial native T1 in COVID-19 survivors (modified/non-linear ShMOLLI colour scale used for purpose of illustration). E, F: Patients had prolonged iron-corrected liver T1 signal, a marker of fibro-inflammation, relative to control. An in-house algorithm was used to calculate iron-corrected T1, so these values cannot be compared to the *LiverMultiScan* cT1[35]. G, H: Renal abnormalities included prolonged kidney T1 and loss of corticomedullary differentiation which were more common in patients recovering from COVID-19 (Units: T2w SI T2-weighted signal intensity; ms milliseconds).

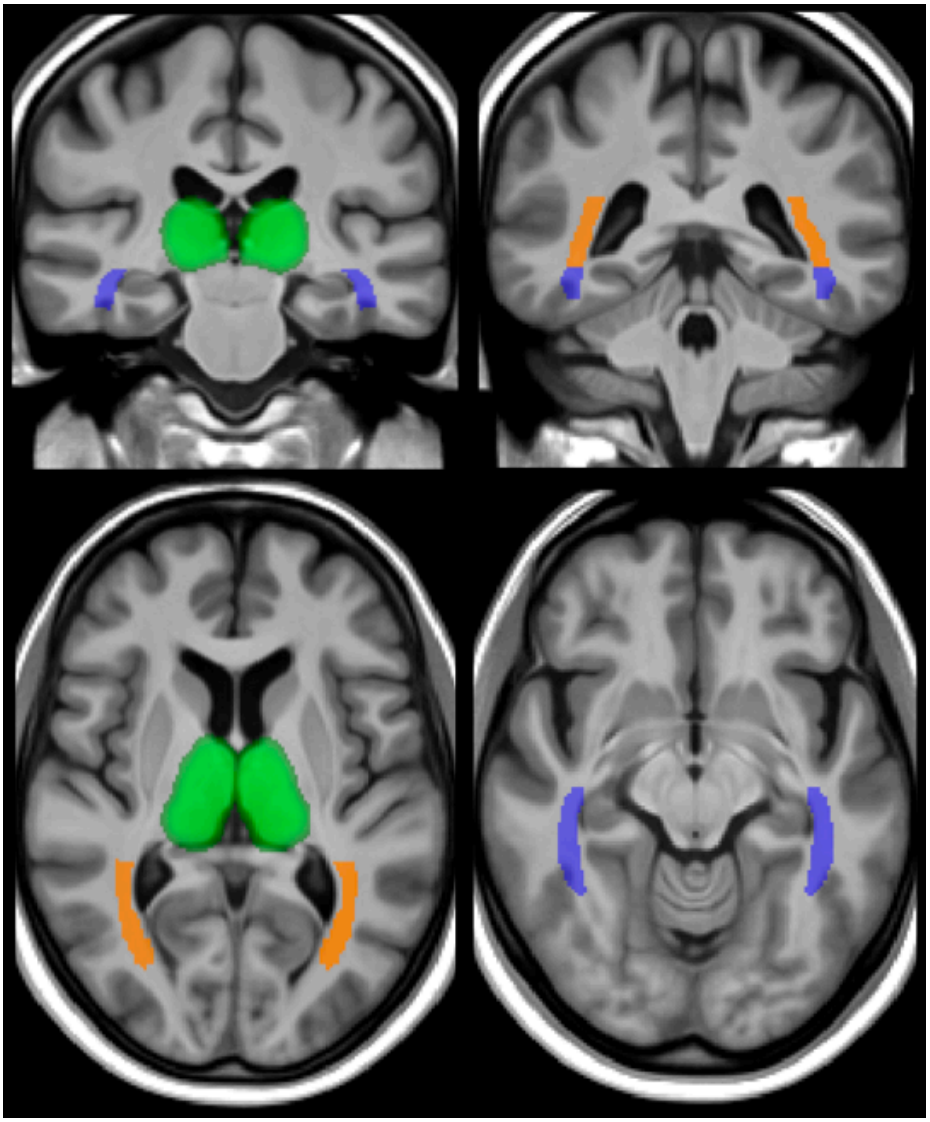

**Appendix Figure 3. Visualisation of brain areas used for extracting imaging-derived phenotypes.**
Green: Thalamus. Orange: Posterior thalamic radiation (includes optic radiation). Blue: Sagittal stratum. The image shows two axial and two coronal sections, with the underlying image being the average T1-weighted structural image from 51 patients recovering from COVID-19, all co-aligned with MMORF[12]
